# Does advance contact with research participants increase response to questionnaires: A Systematic Review and meta-Analysis

**DOI:** 10.1101/2021.02.19.21252094

**Authors:** Benjamin Woolf, Phil Edwards

## Abstract

**Background:** Questionnaires remain one of the most common forms of data collection in epidemiology, psychology and other human-sciences. However, results can be badly affected by non-response. One way to potentially reduce non-response is by sending potential study participants advance communication. The last systematic review to examine the effect of questionnaire pre-notification on response is ten years old, and lacked a risk of bias assessment.

**Objectives:** Update Edwards et al. (2009) to include 1) recently published studies, 2) an assessment of risk of bias.

**Methods:** Data sources: Edwards et al. (2009); 13 data-bases; the references in, and citations of included studies. Eligibility criteria: Randomised control trials examining the impact of pre-notification on response. Data extraction: data extraction was done twice by a single unblinded reviewer. Risk of bias was assessed using the Cochrane Risk of Bias tool and funnel plots.

**Results:** 103 trials were included. Over-all pre-notification increased response, OR = 1.38 (95%CI: 1.25-1.53). However, when studies at high or unclear risk of bias were excluded the effect was greatly reduced (OR = 1.11, 95% CI: 1.01-1.21).

**Conclusions:** The evidence implies that while pre-notification does increase response rates, this may not be of clinical utility.

## Introduction

### Background

Questionnaires have been one of the most common methods of data collection across the social and medical sciences. For example, in epidemiology pen and paper questionnaires alone were used in 29.2% of over 2,000 analytic epidemiological studies included in a review of articles published in high-impact medical journals between 2008 and 2009.^1^ Likewise, about a third of empirical research published in management and accounting journals use questionnaires, and a review of a top social psychology journal found that over 91% of empirical studies published in the second half of 2017 used some form of questionnaire.^2 3^

Inherent in using questionnaires is a risk of non-response. Potential participants, for example, might forget to complete questionnaires, and research ethics requires a right to refuse participation. Non-response can negatively impact on studies in three major ways: Firstly, non-response can introduce selection bias.^4^ Secondly, even in the absence of selection bias, because non-response reduces the number of participants recruited into a study, non-response increases risk of random error (i.e. reduces statistical power and precision). Finally, non-response increases study costs.^5^

It is therefore important to minimise non-response. One potential method is for the study team to contact potential participants in advance of them receiving the questionnaire (questionnaire pre-notification). In 2009, Edwards et al. published a systematic review of randomised control trials evaluating methods of reducing non-response in both postal and electronic questionnaires.^6^ They found that pre-contact increased response when compared to no pre-contact (OR = 1.5, 95% CI 1.26-1.78, for response after first questionnaire administration, and OR = 1.45, 95% CI 1.29-1.63 for response after final questionnaire administration). However, Edwards et al. (2009) did not assess the risk of bias in or across the included studies, and is now 10 years old, so therefore does not include research published in the last decade.

There is therefore a need for an updated review which includes recently published studies, an assessment of bias risk in and across included studies. This review will:

1. Update Edwards et al. (2009)’s systematic review and meta-analysis of randomised control trials examining the effect on non-response of pre-notification relative to no pre-notification (in any population) so that it includes papers published in the last decade.
2. To carry out an assessment of the risk of bias (i) in and (ii) across included studies.

## Methods

### Protocol and registration

The methodology of the review and analysis was approved in advance by the LSHTM epidemiology MSc course directors. A copy of this form, approved on 21/03/2018, can be found in Supplementary Table 1.

This study received ethics approval from the London School of Hygiene and Tropical Medicine MSc Research Ethics Committee on 26/03/2018.

### Eligibility criteria

#### Inclusion criteria

##### Types of population

This study followed Edwards et al. (2009) in using data from “[a]ny population (e.g. patients or healthcare providers and including any participants of non-health studies).” This should maximise generalisability over different contexts.

##### Types of interventions

interventions must include some type of questionnaire pre-contact (pre-notification, advance letter/email/text/phone call or other co-referring term). No restriction is placed on the type of questionnaire pre-notification.

##### Comparison group

Included studies need to be able to make a direct comparison of the effect of questionnaire pre-notification vs no pre-notification (i.e. include at least one arm which received identical treatment to the pre-notification arm other than not receiving the pre-notification).

##### Types of outcome measures

The proportion or number of completed, or partially completed questionnaires returned after all follow-up contacts were complete.

##### Types of study design

Any randomised control trial evaluating a method of advanced contact to increase response to questionnaires. The inclusion of only randomised control trials should on average eliminate risk of confounding biasing estimates within studies.

###### Exclusion criteria

There are no exclusion criteria.

### Information sources

1. Relevant studies identified by Edwards et al. (2009). A detailed description of the information sources, e.g. databases with dates of coverage, used in this study are in its methods section and Supplementary Tables, which can be freely accessed in the Cochrane Library (https://www.cochranelibrary.com/cdsr/doi/10.1002/14651858.MR000008.pub4/full).
2. A search in the same data-bases used in Edwards et al. (2009) from the date they were last searched till the present day. Specifically, the following databases were searched (with date restrictions in brackets): CINAHL (2007.12-2018.6); Dissertation & Thesis, Social Science Citation Index, Science Citation Index, and Index to Scientific & Technical Proceedings in Web of Science (2008.1-2018.6); PsycInfo (2008.1-2018.6); MEDLINE (2007.1-2018.6); EconLit (2008.1-2018.6); EMBASE (2008.1-2018.6); Cochrane Central (2008.1-2018.6); Cochrane CMR (2008.1-2018.6); ERIC (2008.1-2018.6); and Sociological Abstracts (2007.1-2018.6). After consultation with the LSHTM library, two databases searched by Edwards et al. (2009) (National Research Register and Social Psychological Educational Criminological Trials Register) were not searched because they were both deemed inaccessible and no longer operational. Any relevant reviews found in the literature search were examined for relevant studies
3. The references of all included studies, and any citation they, or Edwards et al. (2009), had received by the 28/6/2018 were checked for meeting the eligibility criteria.

Non-English papers were translated using Google Translate.

### Search strategy

The search strategy was developed by modifying the strategy used by Edwards et al. (2009), to make it more sensitive and specific to detecting studies examining questionnaire pre-notification, by adding terms denoting types of pre-notification, and removing terms relating to other methods. The strategy was validated by inputting the new terms into Google Scholar, and checking that it detected all relevant studies included in Edwards et al. (2009). The specific search terms are presenting in Supplementary Table 7.

### Study selection

The eligibility assessment was conducted by one reviewer following a standardised procedure. This process was repeated on a random 10% by a second reviewer with 99.7% agreement. Citations were uploaded onto Covidence (http://www.covidence.org/), a website specially designed for paper screening by the Cochrane Collaboration. Covidence automatically identified duplicates of citation/abstracts, which were then manually checked for errors.

Studies were first screened based on abstracts and titles, then full text. This process was repeated for any study which was referenced by or itself cited by an included study, and on the content of any potentially relevant review identified in the search.

### Data collection process

A standardised data extraction sheet (Supplementary Table 2) was developed. The sheet was pilot tested on 10 randomly chosen studies from Edwards et al. (2009). One reviewer extracted data from included studies. To minimise transcription errors, this process was later duplicated. Authors were contacted for extra information about study bias risk, and still existent copies of communication from Edwards et al. (2009) were examined.

To check for duplication studies which shared at least one author were compared based on similarity of study population, date, and methodology. Duplicate trials were treated as a single study in the meta-analysis.

### Data items

Information extracted for each included trial comprised 5 domains:

1. Information on the inclusion criteria: The study design, nature of the control arm, information on the intervention arm(s), information about the outcome measurement (the number of responses, and/or the response rate, in each arm).
2. Information on risk of bias: how the allocation sequence was generated, information of allocation concealment, blinding of participants and personnel, blinding of outcome assessors, any incomplete outcome data, information on other possible sources of bias (e.g. source of funding).
3. Information on the participants: the total number of participants, numbers in each arm, setting, country.
4. Information on the outcome: number of items returned, or response rate, in each arm.
5. Other information: the time from the sending of pre-notification to questionnaire, if it includes a foot-in-the-door manipulation, the type of questionnaire administration, the type of pre-contact.

### Risk of bias in individual studies

Bias was evaluated using the Cochrane Risk of Bias tool.^7^ Results are stratified based on Risk of Bias score.

### Summary measures

The primary summary measure of association estimated was the ratio of the odds (OR) of response in the treatment groups compared with the odds of response in the control group.

In line with Edwards et al. (2009), the meta-analyses were performed by comparing the ORs using a random-effects model. The analysis was performed on an intention-to-treat basis. Outcomes were only included if they occurred within the period of follow up.

### Synthesis of results/Planned methods of analysis

The results were synthesised in a meta-analysis conducted using STATA 15, using the ‘metan’ command.^8^ To be consistent with Edwards et al. (2009), a random effects meta-analysis was used. Heterogeneity was assessed using the Cochran-Q Chi^2^ statistical test for heterogeneity, and the I^2^ statistic.^9^

### Risk of bias across studies

Risk of bias across studies was assessed with funnel plots. Asymmetry was investigated informally, by visually assessing how symmetrical the plots are around the effect estimate, and formally, using Harbord’s test. Funnel plots were created using the ‘metafunnel’ command in STATA. Because ORs are naturally correlated with their standard error, response rates were used instead of ORs.^8^

## Results

### Study section

A total of 99 papers, reporting a total of 103 trials, were identified for inclusion in the review. The search resulted in a total of 26,894 citations, including 11,408 duplications. The reasons for exclusions are stated in Figure 1 and Supplementary Table 4.The numbers identified and excluded at each stage are described Figure 1.

**Figure 1:**
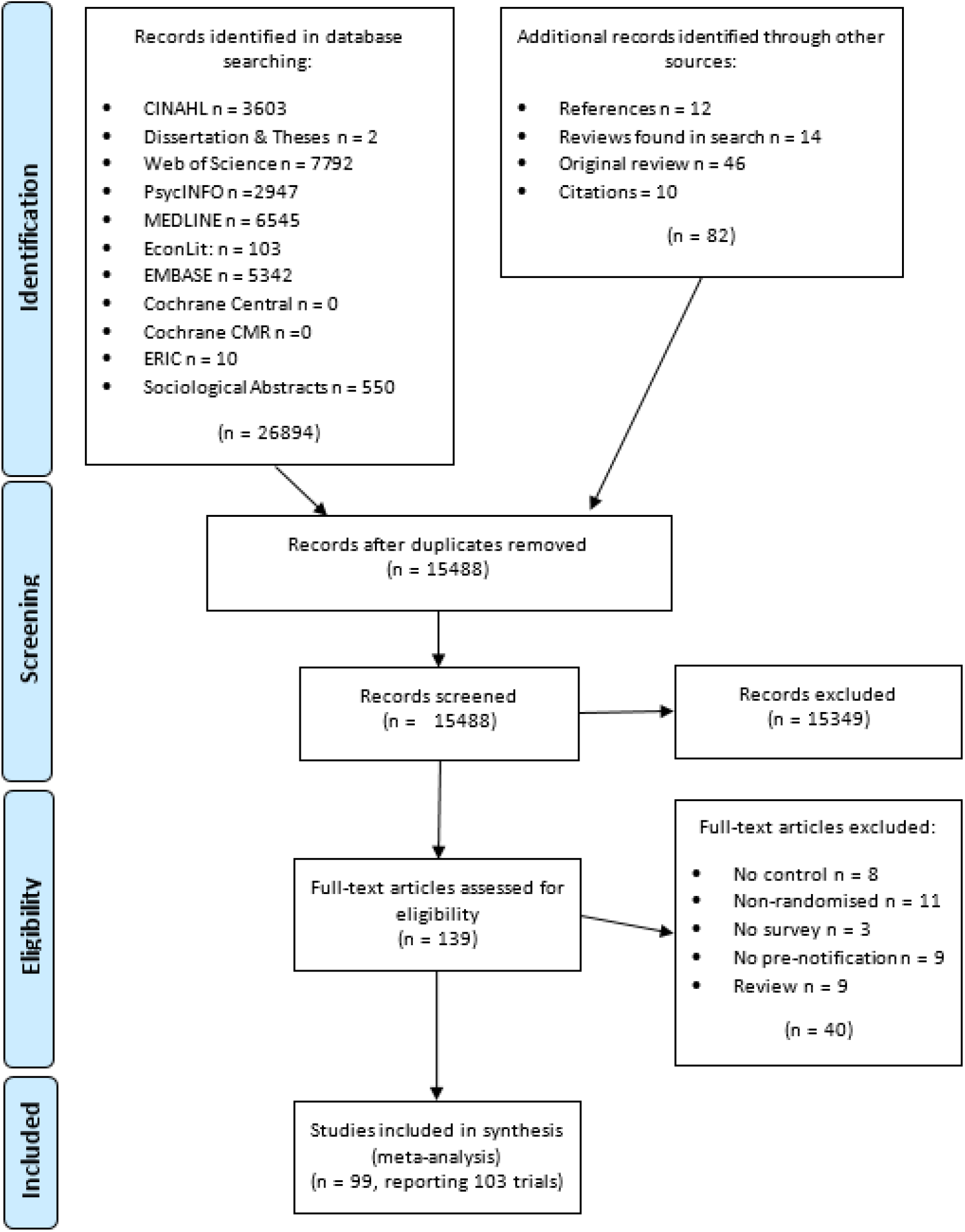
PRISMA flow diagram of information through the different phase of the systematic review

### Study characteristics

The characteristics of the included studies are described in detail in Supplementary Table 5.

### Risk of bias within studies

Judgments formed for each domain of the Cochrane Risk of Bias tool in each study are represented graphically in Figure 2. The supporting evidence can be found in Supplementary Table 6. Overall, 8 studies were at high risk, 8 at low risk and 87 were at unclear risk. The proportions of studies at each level of risk is presented in Figure 3.

**Figure 1:**
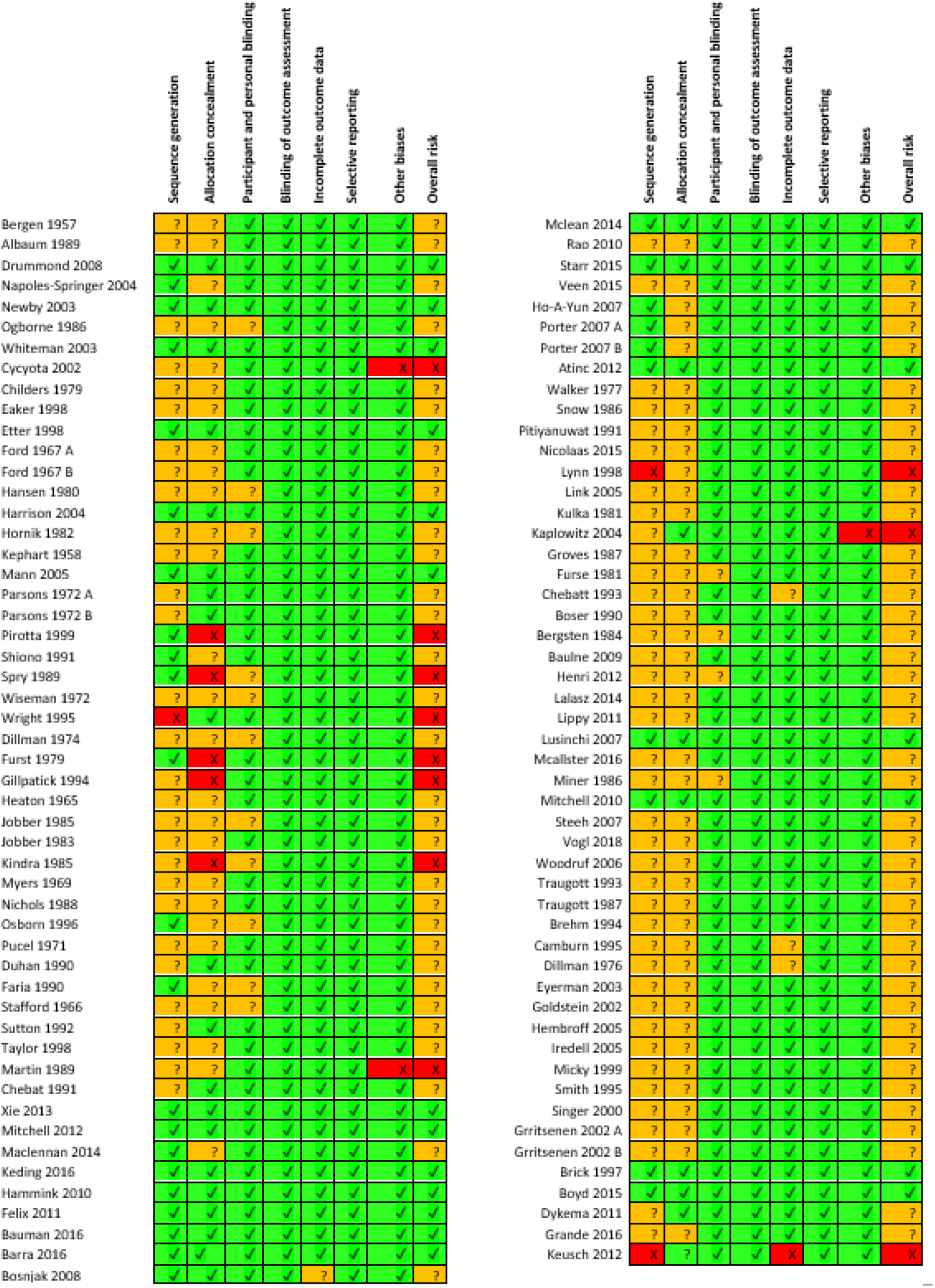
Risk of bias summary figure illustrating judgement about each risk of bias item for each included study.

**Figure 1:**
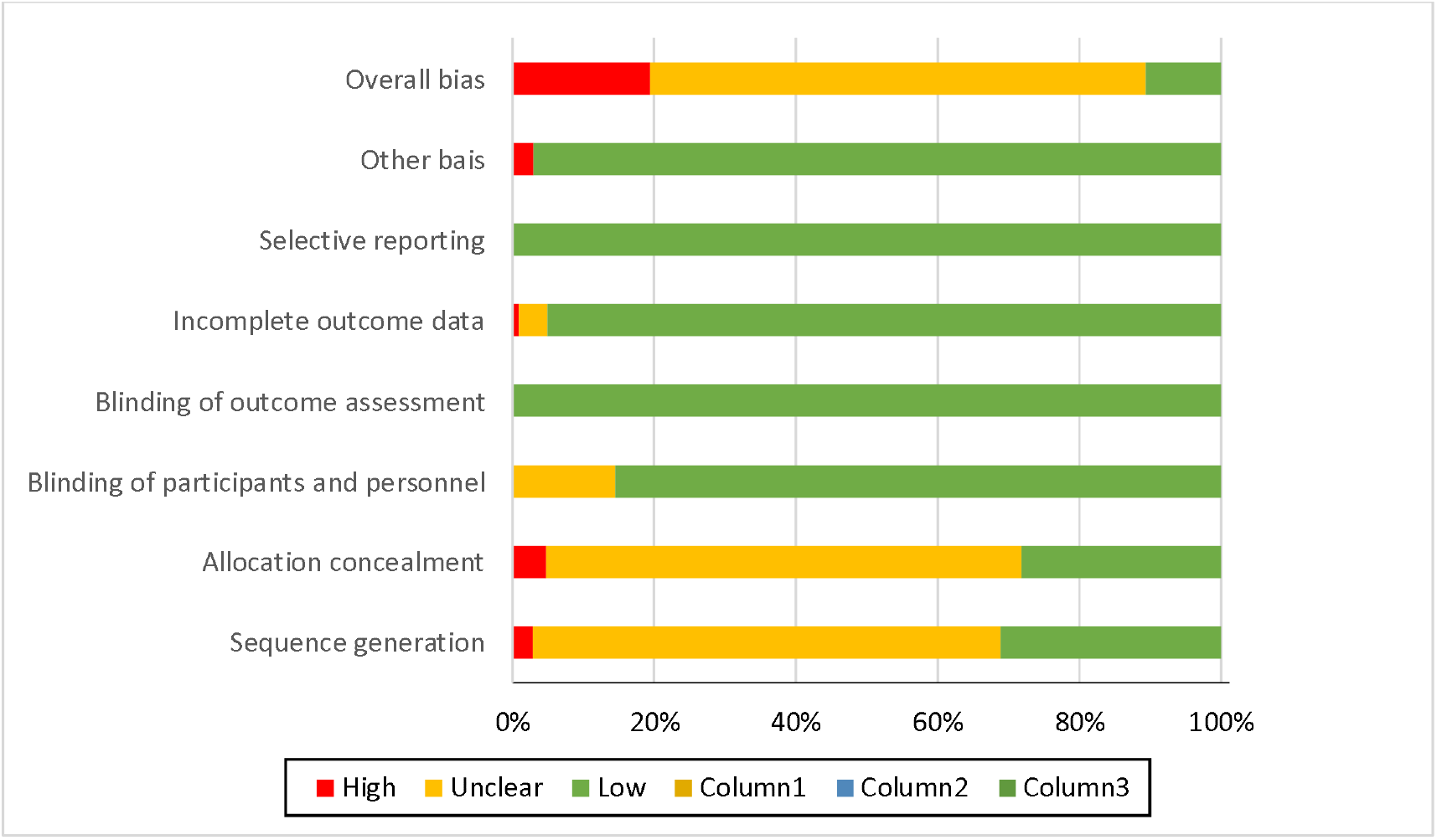
Risk of bias graph illustration judgments about each risk of bias item presented as percentages across all included studies.

### Sequence generation

32 studies described the process used to generate the random sequence, or confirmed the use of randomisation in correspondence. 3 did not use random allocation. 68 studies have an uncertain risk of bias.

#### Allocation concealment

39 studies described concealment, or confirmed it in communication. 5 confirmed that they had not used allocation concealment in communication. The remaining 69 studies provided insufficient information to reach a judgment, and so are of unclear bias.

#### Participant and personnel blinding

Participant and personnel blinding was not reported most trials. However, the design of many trials ensured that a degree of blinding did occur. A common design was to randomise participants to receive or not to receive a pre-notification without prior consent. The pre-notification itself would also often not explain that the participant had been allocated to receive it randomly. Thus any effect of treatment could not be due to the effect of knowing that they had been specially selected for an intervention which others had not got. Although the participant still knew they had received the pre-notification, this knowledge is part of the effect of a pre-notification – and therefore does not introduce any risk of material bias.

Similarly, although most did not describe any blinding procedure for personnel, its absence was often unlikely to lead to bias in estimates. In studies using a pre-written pre-contact (e.g. e-mail, letters, SMS) unblinded study personnel do not have the ability to influence the experience or perceptions of potential participants, as their only means of communication with each other is through a pre-written pro-forma message. This, however, is not true for studies which used a telephone pre-notification, in which the personnel and potential participants can have a genuine interaction. No study with telephone pre-notification reported no blinding of personnel.

Overall 88 studies were regarded as being at low risk of bias, and 15 at unclear risk.

#### Blinding of outcome assessment

Outcome assessment blinding was reported in 8 studies. However, the outcome (whether the questionnaire had been returned) is objective, and unlikely to be influenced by whether the outcome assessor knows the group assignment. Because the analyses are a comparison of two proportions, data analysers were unlikely to have enough researcher degrees of freedom for bias to be introduced in the analyses. All studies were therefore judged as being at low risk of bias for this domain.

#### Incomplete outcome data

98 provided enough information to ascertain the total number of participants randomised in each arm and the total number of questionnaires returned in each arm. However, 4 are at unclear risk because they did not report sufficient detail to estimate per protocol rates, or state if the rates were intention to treat or per protocol, and one study at high risk.

#### Selective reporting

There was little evidence of selective reporting. All studies reported information on the relevant outcomes of interest.

#### Other biases

3 of the factorial studies had significant interaction effects.

### Results of individual studies

The results from individual studies are presented in a forest plot, Figure 4.

**Figure 1:**
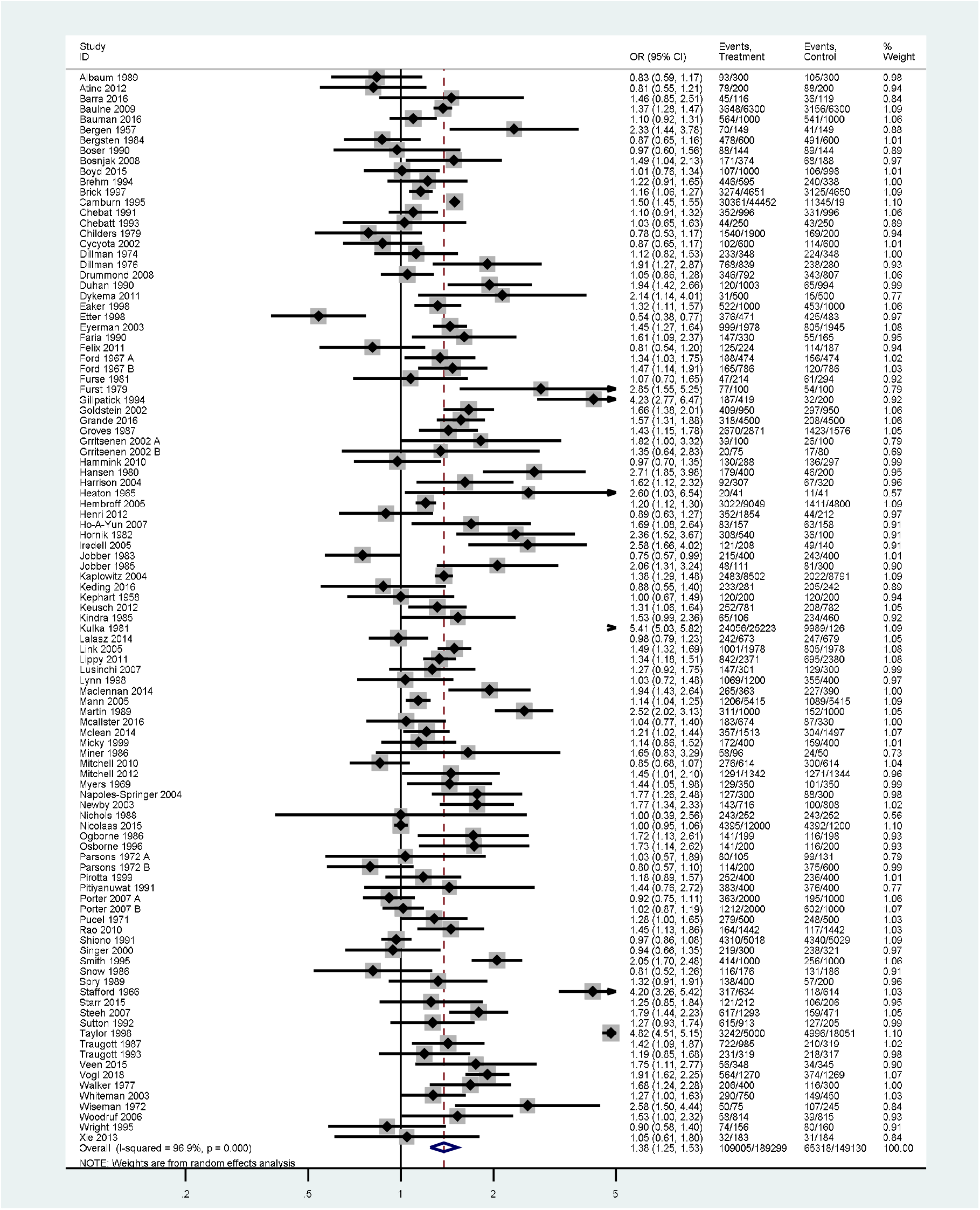
Forest plot of overall response after final follow-up with pre-notification versus no pre-notification.

### Synthesis of results

Information on response was available in all trials, thus data from all trials was used. These randomised a total of 338,429 participants, and had 174,323 returned questionnaires. The pooled estimate shows an increase in response for the final follow-up after questionnaire pre-notification (OR = 1.38, 95% CI: 1.25-1.53, p < 0.001). There was strong evidence of heterogeneity (I^2^ = 96.9%; Tau^2^ = 0.24; *X*^2^ (102, N = 103) = 3311.86, p < 0.001)

### Risk of bias across studies

To explore the possibility of small study bias, funnel plots were created for both outcomes, Figures 5. Visual assessment implies that there is no major asymmetry. However, more studies than expected fell outside the 95% confidence limits. In addition, a formal assessment of asymmetry, using Harbord’s test, did not find evidence to reject the null hypothesis of no asymmetry (p = 0.577).

**Figure 5:**
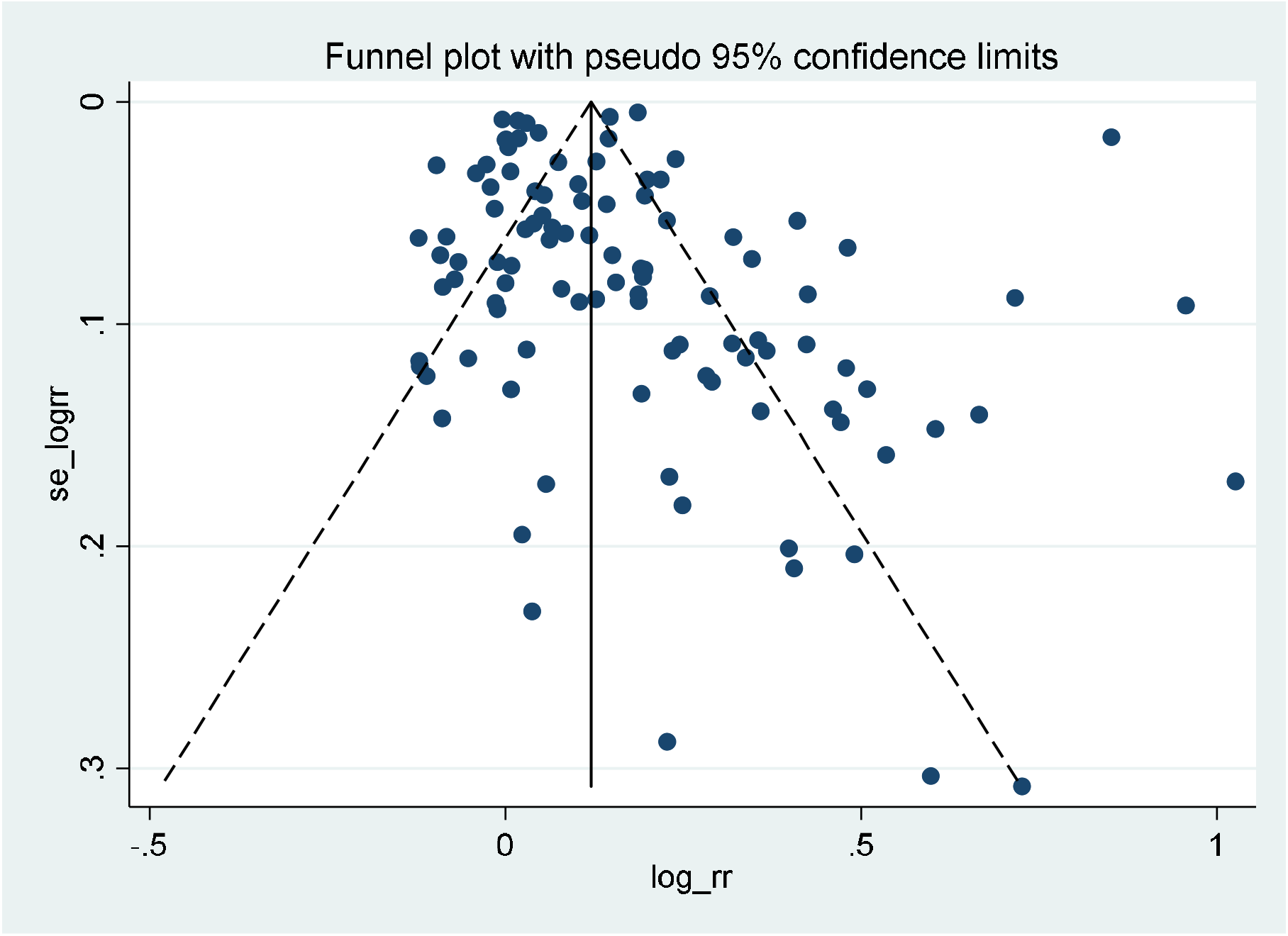
Funnel plot with pseudo 95% confidence limits for response after final follow-up.

### Risk of Bias Within Studies

87 studies were at unclear risk, 8 at low risk, and 8 at high. When stratified by risk of bias, there was no longer evidence against the assumption of pooled association across studies which were of low bias (OR = 1.11, 95% CI: 1.01-1.21 p = 0.027). There was evidence of an association among studies of uncertain risk (OR = 1.45, 95% CI: 1.27-1.66, p < 0.001) and high risk (OR = 1.50, 95% CI: 1.20-1.86, p < 0.001). Heterogeneity was reduced in in both the low (I^2^ = 62.2%; Tau^2^ = 0.02; *X*^2^ (19) = 50.21, p < 0.001) and high (I^2^ = 87.1%; Tau^2^ = 0.11; *X*^2^ (10) = 77.37, p < 0.001) risk groups, but not the unclear risk group (I^2^ = 97.6%; Tau^2^= 0.29; *X*^2^ (71) = 2981.94, p < 0.001). Results displayed in Figure 6.

**Figure 6:**
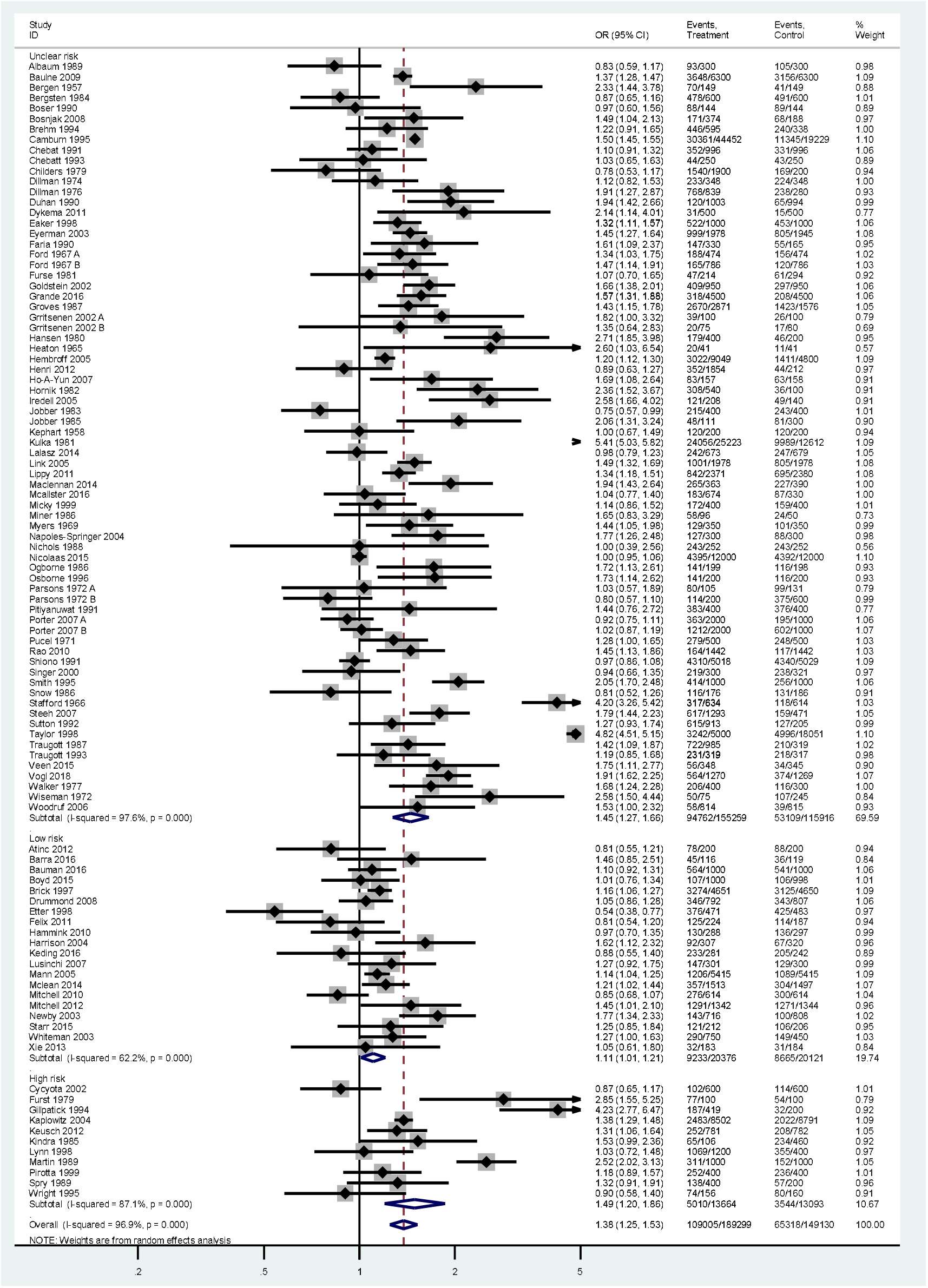
Forest plot of response after final follow-up with pre-notification versus no pre-notification, stratified by risk of bias.

## Discussion and conclusions

### Summary and interpretation of evidence

This meta-analysis and systematic review of randomised control trials examined the effect of pre-notification compared to no pre-notification on questionnaire response rates. Pre-notification led to 1.38 (95% CI: 1.25-1.53) times greater odds for response. However, this was greatly reduced after restricting to studies of low risk of bias, OR = 1.11 (95% CI: 1.01-1.21). This low OR implies that researchers should be cautious when using pre-notification as they may not be cost effective or lead to improvements of clinical relevance.

### Limitations

#### Outcome level limitations

##### Risk of Bias

Across domains, high risk of bias was uncommon. However, few studies provided sufficient information to be assigned low risk of bias. The age of many studies makes communication difficult, e.g. due to address change.

##### Imprecision

Due to the large number of participants in each arm, even after stratification by bias risk, confidence intervals were relatively narrow.

##### Indirectness

There was generally little indirectness in the review. All studies were randomised control trials examining the effect of pre-notification on questionnaire response, so directly answered the study question. However, it is unclear if the study will generalise to any population. For example, there are no studies from low income countries.

##### Publication bias

Visual inspection of the funnel plots and formal testing with Harbord’s test both imply that small study bias was unlikely. As high questionnaire response is important to non-academics, e.g. polling companies, an unassessed grey literature will probably exist.

##### Heterogeneity

There was substantive heterogeneity within the study. Future studies should consider further explanations.

### Review level limitations

#### Search strategy

Cochrane recommends that the literature searching be done by two independent reviewers, while this study only used one.^10^ In addition, the search lacked specificity, and some extra publications might have been found by contacting authors to see if they had published other studies on the question. However, citation searching is not always common in systematic reviews, although it proved an effective way of detecting new studies.

#### Data extraction

Cochrane recommends that data extraction should be done by two independent reviewers, while this study only used one, which should also reduce errors.^11^

### Analysis

### Strengths and weaknesses in relation to other studies

The updated review more than doubled the number of included studies, and four old studies were excluded for poor methodology. The overall results of the two studies are relatively similar, with overlapping confidence intervals overlap the results of the two studies might be consistent. However, restricting to low risk of bias studies implies that this estimate may be due to study bias.

Both studies might be criticised for their choice of outcomes. Response rate does not entail response quality.^5^ For example, a questionnaire might not have been fully completed, or completed inaccurately. In addition, to be a useful intervention for researchers pre-notification needs to be cost effective. However, neither of these outcomes are examined in the reviews.

## Conclusion

This systematic review and meta-analyses of randomised control trials examining the effect of pre-notification on questionnaire response found evidence which supports the use of pre-notification. However, after excluding studies at high or unclear risk of bias the effect of the intervention was greatly reduced, and is probably no longer of clinical relevance.

## Data Availability

All data is available in the supplementary tables.

