## Supplemental Table 1 for "Does advance contact with research participants increase response to questionnaires: A Systematic Review and meta-Analysis"

### Supplementary Table 1: LSTHM Approval Form

**Project title:** The effect of advanced contact with study participants on reducing non-response in research studies.

**Student:** Benjamin Woolf

**Project supervisor:** Phil Edwards (confirmed)

**Personal Tutor:** Anna Goodman

**Type of project (please strike through):**

- Literature review plus quantitative components *(systematic review with meta-analysis)*

**Give an outline of the proposed project, including background to the proposal. *In the following sections* sufficient detail must be given to allow the Course Directors and Ethics Committee to make an informed decision without reference to other documents. If you are proposing a data analysis project *in this background section* please also specify the *sources of secondary data proposed.* If you are proposing a systematic review specify number of eligible studies identified on quick literature search.**

***(300 word limit)***

All research requires people to participate in studies. Non-response by study participants can negatively impact on studies in two major ways. Firstly, if the sample was selected randomly, or assigned an exposure randomly, non-response risks biasing any estimate of effect or prevalence derived from the sample. Secondly, non-response reduces a study’s power to detect a difference.

In 2009 Edwards et al published a systematic review of randomised control trials evaluating different methods of reducing non-response in both postal and electronic questionnaires. This study found that that there was, on average, an effect of pre-contact, and that contact by phone was slightly better than by mail. However, there was substantial heterogeneity in the results: P<0.000001, I^2=91% for the proportion of questionnaires returned after the first mailing, and P < 0.00001 I^2 = 89% for the proportion of questionnaires returned after the final follow-up. In addition, the systematic review is now almost 9 years old, and thus is somewhat out of date.

This project will therefore seek to update Edwards et al (2009)’s systematic review and to assess if the heterogeneity in the results of the effect of pre-contact on response can be explained by of the use of different types of advanced contact. The literature review will include published journal articles only. A preliminary search of studies which cited the approximately 30 studies included in the 2009 review found over 40 potentially relevant studies for this review, of which 10 were published after 2008 and will definitely be included.

**Hypothesis statement**

That the heterogeneity in the study estimates of the effect on response to questionnaires using advanced contact will be reduced if one takes account of the nature of the advanced contact.

**Overall aim of project**

To assess if the type or nature of advanced contact effects survey response.

**Specific objectives of project**

1. Update Edwards et al(2009)’s literature review’s section on examining advanced contact, so that it is up to date.
2. Update Edwards et al(2009)’s meta analyses on advanced contact to include any newer studies
3. Conduct a subgroup analysis to examine any differential effects based on the type of advanced contact, and the extent to which this explains the heterogeneity between studies.

**Specify the procedures/methodology to be conducted during the project. For data analysis projects. *In this methods section include the power available to test your hypothesis and a brief statistical analysis plan*. For literature reviews, include details on search strategy, search terms, inclusion and exclusion criteria.**

Inclusion criteria

Types of studies

Any RCT evaluating any method of advanced contact to increase response to either postal or electronic questionnaires. For example, prior warning by phone/email/post.

Types of data

This study will follow Edwards et al(2009) in using data from “[a]ny population (e.g. patients or healthcare providers and including any participants of non-health studies).”

Types of methods

This study will follow Edwards et al(2009) in including “[a]ny methods designed to increase response to postal or electronic questionnaires. Strategies requiring telephone contact as a follow-up technique are included but those requiring home visits are not.”

Types of outcome measures

This study will use the same outcome measures as examined in Edwards et al (2009).

- Proportion of completed, or partially completed questionnaires returned after the first mailing.
- Proportion of completed, or partially completed questionnaires returned after all mailings.
- Proportion of participants logging-in or clicking the hyperlink to visit the online survey.
- Proportion of participants submitting the online survey.

Search Methods

The following data bases will be searched:

CINAHL 2008-2018.1

ERIC 2008-2018.1

PsycLit 2008.02-2018.1

MEDLINE 2007.12-2018.1

EMBASE 2007.11-2018.1

Social Science Citation Index 2008.2-2018.1

Science Citation Index 2008.2-2018.1

EconLit 2008-2018.1

Sociological Abstracts 2008-2018.1

Index to Scientific & Technical Proceedings 2008.2-2018.1.

((questionnair* OR survey* OR data collection) AND (respon* OR return*) AND (advance* OR earlier OR prior OR before* OR already OR afore OR pre* OR remind* OR communic* OR contact OR notification) AND (control* OR randomi* OR blind* OR mask* OR trial* OR compar* OR experiment* OR “exp” OR factorial))

The following search terms will be used:

A. questionnair* OR survey* OR data collection

B. respon* OR return*

C. advance* OR earlier OR prior OR before* OR already OR afore OR pre* OR remind* OR communic* OR contact OR notification

D. control* OR randomi* OR blind* OR mask* OR trial* OR compar* OR experiment* OR “exp” OR factorial

E. A and B and C and D

We will also search for relevant citations of included studies in Google Scholar.

Data collection and analysis

Trial identification

Myself and Dr Edwards will screen the titles, abstracts and key words of all records identified form the electronic bibliographic databases.

Quality assessment

Trials will be assessed be me on their standard of allocation concealment, blinding, and if they were randomised correctly.

Data extraction

Data will be extracted:

- The nature of advanced contact
- The number randomised to each arm
- the quality of allocation concealment
- The types of participants, materials and follow-up methods used.
- The effect of each intervention on response
- The proportion of questionnaires returned after the first mailing and the proportion returned after all follow up contacts were complete.

**Sub group analyses**

The following subgroups will be examined as prima facie relevant to response:

- Method of contact (e.g. email, telephone, postcard, letter)
- Type of survey (e.g. online, written, telephone)
- Time lag from pre-contact to survey.
- Whether participants could respond to the pre-contact.

**References if appropriate**

Edwards, Philip James, Ian Roberts, Mike J Clarke, Carolyn DiGuiseppi, Reinhard Wentz, Irene Kwan, Rachel Cooper, Lambert M Felix, and Sarah Pratap. “Methods to Increase Response to Postal and Electronic Questionnaires.” In *Cochrane Database of Systematic Reviews*. John Wiley & Sons, Ltd, 2009. http://onlinelibrary.wiley.com/doi/10.1002/14651858.MR000008.pub4/abstract.

**What could stop this project from succeeding, or prevent you from achieving your objectives? Please indicate any aspects of your proposed approach which could potentially experience difficulties, e.g. delays with permissions, data collection or storage problems, lack of sufficient comparable information, etc. You may also wish to mention any wider matters which could affect your project, e.g. civil unrest, natural disasters, transport availability.**

Finding insufficient data to do subgroup analyses.

**What alternative plans do you have (ie what project you could do instead) in case you encounter any of the potential problems you have identified?**

Widen the scope to a more general update of the 2009 systematic review.

**What specific facilities or resources will you personally expect to make use of for your project (eg a local university library, lab facilities, project placement with a specific organisation etc)?**

LSHTM and/or Senate House Library.

**Where will the project be carried out? Note that work away from LSHTM or outside the UK means any form of work for your project, not just primary data collection. *For MSc Epidemiology projects there is insufficient time to do new field work. You will not be permitted to collect any new data unless specific permission from the course directors is sought and granted.***

In LSHTM

**Where the research is to take place overseas, ethical approval must be obtained in the country(s) concerned. Approval from the LSHTM Committee is dependent on local approval having been received. For all countries listed, detail arrangements being made to obtain local ethical and/or regulatory approval. *For the main ethics application (part of the web-based CARE form on LSHTM Ethics online –the LEO system) you will need to electronically append copies of local approval letter(s) where this has already been obtained*. Where you believe local approval is not required, please explain why not and describe any less formal permissions, invitations or support you are being given for this work.**

na

**Where the research is taking place in the UK, please list other UK Committees from which approval is being sought*. For the online CARE form (main ethics application) you will need to electronically append copies of local approval letter(s) where this has already been obtained.***

This research will only take place at LSHTM.

**If you expect to use any public domain data, please give further details. Make clear who owns the data and how you will gain access (giving a link if possible). Public domain data must be available to any member of the public, without any restrictions or requirement for special permission, and must not enable the identification of living people.**

The project will extract summary statistics form published papers. Some of this might be in the public domain, however it should not allow for identifiability.

**If you expect to use existing data, how will you obtain it? Indicate who holds the data, who specifically you will contact, and by when. Any contact so far, especially anything confirmed in writing, should be mentioned.**

Due to time limitations this project will only use data extractable from available published journal articles – using the methods described above. No advanced contact has been made with paper’s authors, however, this should be unnecessary to extract the required information.

**Will any copyright agreements or intellectual property rights (IPR) agreements be required regarding data to be used or collected in the project? Please tick all boxes that apply. For the main ethics application you may be asked to provide copies of any forms/agreements (even if in draft).**

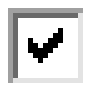
No specific IPR, copyright or permissions issues apply to this project (student retains copyright & claim to related IPR)

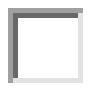
IPR to be retained by LSHTM (specific LSHTM form to be completed)

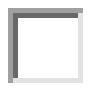
Copyright to be transferred to LSHTM (specific LSHTM form to be completed)

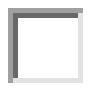
IPR, copyright or other agreements/permissions required with external parties/organisations
