## Supplemental Table 2 for "Does advance contact with research participants increase response to questionnaires: A Systematic Review and meta-Analysis"

Supplementary Table 2: Data Extraction Form

| ID |
| --- |
| Intervention Arm(s) |
| Control Arm(s) |
| Outcome definition |
| Number in control arm(s) |
| Number in intervention arm(s) |
| Number/ rate of return in control arm |
| Number /rate of return in intervention arm |
| Total number of Participants |
| Length of data-collection period |
| Setting |
| Country |
| Questionnaire topic |
| Other design comments |
| Delay form pre-contact to questionnaire |
| Method of per-notification |
| Method of sending questionnaire |
| Foot-in-the-door? |
| Sequence generation |
| Allocation confinement |
| Participant and personnel blinding |
| Blinding of outcome assessment |
| Incomplete outcome data |
| Selective reporting |
| Other bias |
