## Supplemental Table 3 for "Does advance contact with research participants increase response to questionnaires: A Systematic Review and meta-Analysis"

### Supplementary Table 3: Reviews found in the literature search and examined for relevant studies

Edwards, P., Roberts, I., Clarke, M., DiGuiseppi, C., Pratap, S., Wentz, R., Kwan, I. and Cooper, R., 2007. Methods to increase response rates to postal questionnaires. *Cochrane database of systematic reviews*, (2).

Edwards, P.J., Roberts, I., Clarke, M.J., DiGuiseppi, C., Wentz, R., Kwan, I., Cooper, R., Felix, L.M. and Pratap, S., 2009. Methods to increase response to postal and electronic questionnaires. *The Cochrane Library*.

Lacey, R.J., Wilkie, R., Wynne-Jones, G., Jordan, J.L., Wersocki, E. and McBeth, J., 2017. Evidence for strategies that improve recruitment and retention of adults aged 65 years and over in randomised trials and observational studies: a systematic review. *Age and ageing*, *46*(6), pp.895-903.

Martins, Y., Lederman, R.I., Lowenstein, C.L., Joffe, S., Neville, B.A., Hastings, B.T. and Abel, G.A., 2012. Increasing response rates from physicians in oncology research: a structured literature review and data from a recent physician survey. *British Journal of Cancer*, *106*(6), p.1021.

Treweek, S., Mitchell, E., Pitkethly, M., Cook, J., Kjeldstrøm, M., Johansen, M., Taskila, T.K., Sullivan, F., Wilson, S., Jackson, C. and Jones, R., 2010. Strategies to improve recruitment to randomised controlled trials. *The Cochrane Library*.

Treweek, S., Pitkethly, M., Cook, J., Fraser, C., Mitchell, E., Sullivan, F., Jackson, C., Taskila, T.K. and Gardner, H., 2018. Strategies to improve recruitment to randomised trials. *The Cochrane Library*.

van Gelder, M.M., Vlenterie, R., IntHout, J., Engelen, L.J., Vrieling, A. and van de Belt, T.H., 2018. Most response-inducing strategies do not increase participation in observational studies: a systematic review and meta-analysis. *Journal of clinical epidemiology*, *99*, pp.1-13.

Weiner, M.D., Puniello, O.T. and Noland, R.B., 2016. Conducting Efficient Transit Surveys of Households Surrounding Transit-Oriented Developments. *Transportation Research Record: Journal of the Transportation Research Board*, (2594), pp.44-50.

Young, T. and Hopewell, S., 2011. Methods for obtaining unpublished data. *The Cochrane database of systematic reviews*, (11), pp.MR000027-MR000027.
