## Supplemental Table 4 for "Does advance contact with research participants increase response to questionnaires: A Systematic Review and meta-Analysis"

### Supplementary Table 4: studies excluded

| **Study ID** | **Reason for Exclusion** |
| --- | --- |
| Beebe 2010 | No Control group |
| Bhutta 2013 | No Randomisation |
| Duncan 2013 | No Questionnaire |
| Dykema 2013 | No Randomisation |
| Edelman 2013 | No Pre-Notification |
| Edwards 2016 | No Randomisation |
| Edwards 2007 | Review |
| Edwards 2009 | Review |
| Gattellari 2012 | No Control Group |
| Grava-Gubins 2008 | No Pre-Notification |
| Green 2015 | No Randomisation |
| Greenfield 2012 | No Pre-Notification |
| Hales 2018 | No Pre-Notification |
| Hoisak 2014 | No Pre-Notification |
| Jacob 2012 | No Control Group |
| Koitsalu 2018 | No Randomisation |
| Koopman 2013 | No Randomisation |
| Lacey 2017 | Review |
| Leathem 2009 | No Pre-Notification |
| Libby 2011 | No Questionnaire |
| Lynn 2016 | No Control Group |
| Mann 2015 | No Control Group |
| Martins 2012 | Review |
| Millar 2011 | No Pre-Notification |
| O’Carroll 2015 | No Control Group |
| Senore 2015 | No Questionnaire |
| Slater 2016 | No Pre-Notification |
| Todd 2015 | No Control Group |
| Treweek 2018 | Review |
| Treweek 2010 | Review |
| Van Gelder 2018 | Review |
| Wagner 2017 | No Randomisation |
| Weiner 2016 | Review |
| Westrick 2017 | No Pre-Notification |
| Young 2011 | Review |
| Scott 1958 | No Randomisation |
| Temple –Smith 1998 | No Control Group |
| Wynn 1985 | No Randomisation |
| Waisanen 1954 | No Randomisation |
| Murphy 1991 | No Randomisation |
