## Supplemental Table 5 for "Does advance contact with research participants increase response to questionnaires: A Systematic Review and meta-Analysis"

### Supplementary Table 5: Full summary of included studies evaluating the effect of pre-notification on questionnaire response

| **Citation** | **Comparison** | **Outcome definition** | **Design** | **Setting (Country)** | **Topic** | **Delay**  **Length** | **Pre-Contact Method** | **Survey Delivery** | **Foot-in-the-door?** |
| --- | --- | --- | --- | --- | --- | --- | --- | --- | --- |
| Bergen 1957 | Pre-contact or Control | final follow-up | experiment | primary school teachers  (Netherlands) | options and attitudes towards public opinion researcher |  | mail | Postal | have to return a pre-paid return card |
| Albaum 1989 | (Pre-contact or Control)x(leaflet or control) | final follow-up | factorial experiment | Business firms who do international market activities. (Denmark) | questions about work |  | mail | Postal | send description |
| Drummond 2008 | (Pre-contact vs Control) x(questionnaire or control) | final follow-up | factorial experiment | GPs. (Ireland) | Health: views and practices about prostate-specific testing (PSA) | 3 weeks | mail | Postal | none stated |
| Napoles-Springer 2004 | Pre-contact or Control | final follow-up | Randomised control trail. | Nested in satisfaction survey of ambulatory care clinics. Have to use primary care and be older than 50 (USA) | about hospital experience year before/stratification | 2 weeks | mail | Postal | none stated |
| Newby 2003 | Pre-contact, Control, colour follow up, or monetary incentive | first and final follow-up | Randomised control trail. | random sample of business in Perth. exclude gov. enterprises and publicly owned firms (Australia) | about business: expectations and attitudes of the self employed | 2 weeks | telephone | Postal | Asked relevant questions |
| Ogborne 1986 | Pre-contact or Control | first and final follow-up | Randomised control trail. | health and social workers (Canada) | about work |  | telephone | Postal (phone option in intervention) | offer a telephone interview if better than mail |
| Whiteman 2003 | x2 types of incentives, Pre-contact or Control | first and final follow-up | Randomised control trail. | women age 40-60 and in Baltimore (USA) | women’s health | 1 week | mail | Postal | none stated |
| Cycyota 2002 | (Pre-contact or Control) x(incentive or cont.) x (personalisation or cont. ) x(follow-up or cont.) x (postage or cont.) | final follow-up | factorial experiment | chamber of commerce survey to business (USA) | business climate | 2 weeks | mail | Postal | none stated |
| Childers 1979 | x2 types Pre-contact or Control | final follow-up | Randomised control trail. | agents of a large Midwest-based insurance company (USA) | insurance |  | mail | Postal | one group given return cards |
| Eaker 1998 | (Pre-contact or Control) x (length or control) x (mention of telephone contact or control) | final follow-up | factorial experiment | Men and women living in Sweden in 1995 20-79 yrs. old | health risk factors | 1 week | mail | Postal | none stated |
| Etter 1998 | (Pre-contact or control) x (layout or control) | first and final follow-up | factorial experiment | annual insurance questionnaire; residents of Geneva, valid address | health insurance survey | 2 weeks | mail | Postal | none stated |
| Ford 1967 A | Pre-contact or control | first and final follow-up | experimental design | Residents of Chenoa (USA) | shopping survey | 1 week | mail | Postal | none stated |
| Ford 1967 B | Pre-contact or control | first and final follow-up | experimental design | Residents of Beardstown (USA) | shopping survey | 1 week | mail | Postal | none stated |
| Hansen 1980 | x2 Pre-contact, questionnaire length, or control | final follow-up | Randomised control trail. | People who bought cars in past year in Ohio (USA) | consumer’s attitudes towards recent new car purchases | 3 days | telephone | Postal | Asked if willing to enter study. |
| Harrison 2004 | Pre-contact or control | final follow-up | Randomised control trail. | patients referred to exercise referral scheme in past 12 months by primary care | survey on relation between service expectations and outcomes | 1 week | mail | Postal | none stated |
| Hornik 1982 | Pre-contact or control | final follow-up | Randomised control trail. | Sample from telephone directly (USA) | about TV/advertising | under 1 week | telephone | Postal | none stated |
| Kephart 1958 | Pre-contact or control | first and final follow-up | experiment | women who had taken state nursing exam in 1950 (USA) | Attitudes towards nursing profession | 1 week | mail | Postal | none stated |
| Mann 2005 | Pre-contact or control | final follow-up | experiment | registered voters in 3 states (USA) | election survey |  | mail | telephone | none stated |
| Parsons 1972 A | Pre-contact or control | only one mailing | experiment | MBA alumni (USA) | politics and religion | 4 days | mail | Postal | none stated |
| Parsons 1972 B | Pre-contact or control | only one mailing | experiment | leaders of 2 religious sects (USA) | politics and religion | 5 days | mail | Postal | none stated |
| Pirotta 1999 | Pre-contact or control | first and final follow-up | Randomised control trail. | GPs. From health insurance In Victoria; have to have had 1500 consultations in prior year (Australia) | work | 5 days | mail | Postal | ask for prompt return |
| Shiono 1991 | Pre-contact or control | first and final follow-up | Randomised control trail. | physician who graduated from med school in 1985 (USA) | survey of pregnancy in physicians. Mailed was personalised | 1 week | mail | Postal | toll free phone number to call if any questions about the survey |
| Spry 1989 | Pre-contact or control x 3 | first and final follow-up | factorial experiment | residence of San Diego (USA) | survey on health related behaviour. Enrolments from medical school director | under 1 week | phone or post card | Postal | none stated |
| Wiseman 1972 | 2 types of Pre-contact or control | final follow-up | experiment | residents of Boston (USA) | political issue polling | under 1 week | Telephone or mail | Postal | describes survey |
| Wright 1995 | Pre-contact or control | first and final follow-up | experiment | census of local government representatives (USA) | commercial census | 2 weeks | mail | Postal | Asked if unwilling to enter study. |
| Dillman 1974 | Pre-contact or control | final follow-up | Randomised control trail. | sample of general public (USA) | feelings and concerns about Washington State University | na | telephone | Postal | ask questions to raise salience |
| Furst 1979 | (Pre-contact or control) x (Personalisation or contorl) | final follow-up | factorial experiment | head teachers (USA) | personality test | under 1 week | mailed | Postal | none stated |
| Gillpatick 1994 | (Pre-contact or control) x (2 types of monetary incentive or control) | only one mailing | factorial experiment | engineers who subscribe to a trade journal (USA) | market research (how good a CAD program is). One condition is personally pre-contacted, the other gets a referral from a colleague |  |  | Postal | none stated |
| Heaton 1965 | Pre-contact or control | only one mailing | experiment | people who bought a Chevrolet in Philadelphia | car sales survey. Attempt to show importance of survey. Also personalised (e.g. hadn’t signed) | 1 week | mailed | Postal | none stated |
| Jobber 1985 | Pre-contact or control | final follow-up | Randomised control trail. | UK textile companies executives | Explore the design and extent of implementation of marketing information system |  | telephone | Postal | none stated |
| Jobber 1983 | (Pre-contact or control) x (colour or control) | first and final follow-up | factorial experiment | UK textile companies | marketing practices |  | mailed | Postal | none mentioned |
| Kindra 1985 | (Pre-contact or control) x (incentive or control) | first and final follow-up | factorial experiment | telephone directory (Canada) | response to advertising |  | telephone | Postal | Asked questions in pre-contact |
| Myers 1969 | follow up, Pre-contact or control | only one mailing | Randomised control trail. | telephone directory (USA) | reaction to bank advertisement | 1 week | mailed | Postal | none stated |
| Nichols 1988 | Pre-contact or control) | final follow-up | Randomised control trail. | sample of electoral role (UK) | mailed was a leaflet on nutrition + cover mailed | 5 weeks | mailed | Postal | none stated |
| Osborn 1996 | Pre-contact or control | first and final follow-up | Randomised control trail. | GPs (Australia) | view on pathology test |  | telephone | Postal | none stated |
| Pucel 1971 | Pre-contact or control | only one mailing | Randomised control trail. | graduates from 24 post-high schools (USA) | effect of training | 1 week | mailed | Postal | none stated |
| Duhan 1990 | Pre-contact or control | first and final follow-up | Randomised control trail. | marketing executives (USA) | work related | 1 week | mailed | Postal | Asked questions |
| Faria 1990 | x2 types of Pre-contact or control | first and final follow-up | Randomised control trail. | Homeowners residing on the property owners’ listing (USA) |  | under 1 week | phone or mailed | Postal | Asked if they will participate |
| Stafford 1966 | x2 types of Pre-contact or control | only one mailing | Randomised control trail. | students (USA) | collegiate clothing | under 1 week | phone or mailed | Postal | none stated |
| Sutton 1992 | (personalisation or control) x (Pre-contact or control) | first and final follow-up | factorial experiment | customers of a utility company, and contractors (USA) | reaction to an established energy rebate program | 10-14 days: phone, 1 week: card | mailed | Postal | none stated |
| Taylor 1998 | Pre-contact or control | first and final follow-up | Randomised control trail. | Young people in the Youth Cohort Study 8 sample, (UK) | Attitudes and behaviour | 1 week | mailed | Postal | none stated |
| Martin 1989 | (Pre-contact or control) x (follow up or control) x (personalisation or control) x ( cover mailed or control) x (return postage or control) | final follow-up | factorial experiment | students in an urban university (USA) | views on the university |  | mailed | postal | none stated |
| Chebat 1991 | (Pre-contact or control) x (incentive or control) | final follow-up | factorial experiment | The Quebec population within the legal driving age (Canada) |  |  |  | Postal | none stated |
| Xie 2013 | Pre-contact or control | final follow-up | Randomised control trail. | female nurses, age 35-65. with correct contact information (Hong Kong) | work and health | 1 week | mail | Postal | asked to send reply slip |
| Mitchell 2012 | Pre-contact or control | final follow-up | Randomised control trail. | nested in follow up of the SCOOP clinical trial. Women aged between 70-84, at high risk of osteoporotic features (UK) | Trail questions | 6 weeks | mail | Postal | none stated |
| Maclennan 2014 | Pre-contact or control | first and final follow-up | Randomised control trail. | nested in RECORD clinical trial. Patients who had not responded to annual follow ups. over 70, history of fracture, not in other methodological study (UK) | self-reported fracture and quality of life | ≥ two weeks | telephone | Postal | none stated |
| Keding 2016 | Pre-contact or control | only one mailing | Randomised control trail. | nested in ACUDep trial, primary care patients in N England. Have mobile phone | quality of life | 4 days | sms | Postal | none stated |
| Hammink 2010 | (Pre-contact or control) x (follow up or control) | first and final follow-up | factorial experiment | nested in survey of GP patients (Netherlands) | quality of care/experience | 1 week | mail | Postal | none stated |
| Felix 2011 | (Pre-contact or control) x (tone or control) | only one mailing | factorial experiment | authors of published maternal health research (UK) | applying their research to LIC | 1 week | email | online | none stated |
| Bauman 2016 | (Pre-contact or control) x (follow up or control) | first and final follow-up | factorial experiment | nested in 45 and Up Study. adults 45 to 100 living in New South Wales. (Australia) | socio-environmental causes of health | 2 weeks | mail | Postal | none stated |
| Barra 2016 | Pre-contact or control | first and final follow-up | Randomised control trail. | patients discharged in stated time who had not responded to previous survey, involved in other studies/care and had a phone number (Norway) | post stroke questionnaire | 1 week | mail | Postal | ask for consent to receive survey |
| Bosnjak 2008 | (x2 types Pre-contact or control) x (invitation or control) | final follow-up | factorial experiment | university students (Germany) | psychometrics, e.g. personality test | 1 week | email or sms | online | none stated |
| Boyd 2015 | (Pre-contact or control) x2 follow up or control) x (design or control) | final follow-up | factorial experiment | ALSPAC follow up (UK) | consent to patients in follow up. | 1 week | mail | Postal | none stated |
| Dykema 2011 | (Pre-contact or control) x (incentive or control) | final follow-up | factorial experiment | physicians, (USA) | assess knowledge of genetic variation. | 1 week | mail | online | none stated |
| Grande 2016 | Pre-contact or control | only one mailing | Randomised control trail. | Random digit dialling (Australia) | epidemiological facts about the workplace | same day | sms | phone | none stated |
| Keusch 2012 | (Pre-contact or control) x (senders gender or control) x (design or control) | only one mailing | factorial experiment | IT managers (Austria) |  | 1 week | email | online | none stated |
| Mclean 2014 | (Pre-contact or control) x (design or control) | only one mailing | factorial experiment | the electoral roll (Australia) |  | 1 week | mail | Postal | none stated |
| Rao 2010 | (Pre-contact or control) x (follow up or control) x (incentive or control) x (recurrent mode or control) | final follow-up | factorial experiment | Random digit dialling | opinion poll survey | 1 week | mail | Postal | none stated |
| Starr 2015 | (Pre-contact or control) x (follow up or control) | final follow-up | factorial experiment | nested in epilepsy rct, provided phone number (UK) |  | under a week | sms | Postal | none stated |
| Veen 2015 | X2 types Pre-contact or control | only one mailing | Randomised control trail. | university students (Germany) | cheating in exams | 1 week | mail | online | none stated |
| Ho-A-Yun 2007 | Pre-contact or control | only one mailing | Randomised control trail. | GPs in Scotland (UK) |  | under a week | telephone | Postal | none stated but uses phone to call receptionist |
| Porter 2007 A | (x2 types Pre-contact or control) x (follow up or control) | final follow-up | factorial experiment | high school students who contacted liberal arts college but did not apply (USA) | perceptions of college | 1 week | email or post | online | none stated |
| Porter 2007 B | (x2 types Pre-contact or control) x (follow up or control) | final follow-up | factorial experiment | alumni of liberal arts collage (USA) | career post-graduation | 1 week | email or post | online | none stated |
| Atinc 2012 | (Pre-contact or control) x (follow up or control) | final follow-up | factorial experiment | university staff and faculty (USA) |  | under a week | email | online | none stated |
| Walker 1977 | Pre-contact or control | only one mailing | Randomised control trail. | credit card holders (USA) | consumer credit survey/purchase history | under a week | mailed | mail | none stated |
| Snow 1986 | Pre-contact or control | final follow-up | Randomised control trail. | People existing job training partnership program (USA) | outcome of state-wide job training program | 1.5weeks | mailed | phone | none stated |
| Pitiyanuwat 1991 | (Pre-contact or control) x (deadline or control) x (design or control) | first and final follow-up | factorial experiment | public school teachers, (Thailand) | desirable characteristics of a teacher |  | mailed | mail | none stated |
| Nicolaas 2015 | (Pre-contact or control) x (follow up or control) x (design or control) | final follow-up | factorial experiment | embedded in a GP patient survey: over 18, registered with GP for >6mths (UK) | expense of patients of the NHS | 1 week | mailed | mail | none stated |
| Lynn 1998 | Pre-contact or control | only one mailing | Randomised control trail. | BT communication survey (UK) | Communication habits of GB population. |  | mailed | mail | none stated |
| Link 2005 | Pre-contact or control | only one mailing | Randomised control trail. | House holds in Behaviour Risk Factor Survey (USA) | health behaviours | under a week | mailed | phone | none stated |
| Kulka 1981 | (Pre-contact or control) x (incentive or control) x (extra follow up or control) x (postage or control) | final follow-up | factorial experiment | registered nurses enrolled in a survey (USA) |  | 2 weeks | mailed | mail | none stated |
| Kaplowitz 2004 | Pre-contact or control | final follow-up | Randomised control trail. | university students (USA) |  |  | mailed | email | none stated |
| Groves 1987 | X3 types of Pre-contact or control | only one mailing | Randomised control trail. | nested in NHIS survey (USA) | Health care |  | mailed | phone | none stated |
| Furse 1981 | Pre-contact, incentive or control | final follow-up | Randomised control trail. | Tennessee population (USA) |  |  | phone | mail | none stated |
| Chebatt 1993 | (questionnaire type or control) x (Pre-contact, incentive or control) | first and final follow-up | factorial experiment | (Canada) |  | 2 weeks | mailed | mail | none stated |
| Boser 1990 | (Pre-contact or control) x (follow up or control) | final follow-up | factorial experiment | Graduates (USA) | emphasise value of participation | 1 week | mailed | mail | none stated |
| Bergsten 1984 | Pre-contact or control | only one mailing | Randomised control trail. | Medicare beneficiaries 65+ (USA) | access to health care | 1 week | phone | interview | arrange time for interview |
| Baulne 2009 | Pre-contact or control) | only one mailing | Randomised control trail. | people over 15 and live in a household. (Canada) | health |  | mail | phone | none stated |
| Henri 2012 | X2 types Pre-contact or control | final follow-up | Randomised control trail. | listed companies (Canada) | management accounting research | 2 weeks | telephone or mail | mail | none stated |
| Lalasz 2014 | Pre-contact or control | final follow-up | Randomised control trail. | alumni 1 year post graduation (USA) | graduate careers etc. | 2 weeks | mail | online | none stated |
| Lippy 2011 | Pre-contact or control | only one mailing | Randomised control trail. | nested in German Health Update Survey 2009. | health | 2 weeks | mail | phone | none stated |
| Lusinchi 2007 | Pre-contact or control | first and final follow-up | Randomised control trail. | electrical engineers (USA) |  | under a week | email | online | none stated |
| McAllster 2016 | X2 types Pre-contact or control | first and final follow-up | Randomised control trail. | university staff (USA) |  | under a week | post or email | mail | none stated |
| Miner 1986 | X2 types Pre-contact or control | final follow-up | Randomised control trail. | parents/carers of people who had used a child psychiatry unit (USA) |  | under a week | post or phone | mail | none stated |
| Mitchell 2010 | (Pre-contact or control) x (follow up or control) | only one mailing | factorial experiment | Academics in Northern UK |  | 6 weeks | mail | mail | none stated |
| Steeh 2007 | sms + called back (passive), sms+ user has to call (active) or control | only one mailing | Randomised control trail. | Nexel subscribers. (USA) |  | Under 1 week | sms | phone | one treatment group have to call rather than be called to do interview |
| Vogl 2018 | Pre-contact or control | only one mailing | Randomised control trail. | Random digit dialling (Germany) | Partner violence | 1 week | mail | phone | none stated |
| Woodruf 2006 | Pre-contact or control | final follow-up | Randomised control trail. | Random digit dialling (USA) | info about study/importance of patients. Parents/children for NIH study on teenage health | 2 weeks | mail | phone | none stated |
| Traugott 1993 | Pre-contact or control | only one mailing | Randomised control trail. | Random digit dialling (USA) |  |  | mailed | phone | none stated |
| Traugott 1987 | (Pre-contact or control) x (personalisation or control) | only one mailing | factorial experiment | Random digit dialling (USA) |  |  | mailed | phone | none stated |
| Brehm 1994 | inactive + Pre-contact, Pre-contact, logo, or control | only one mailing | Randomised control trail. | Random digit dialling (USA) | Attitudes to war | 1-2 weeks | mailed | phone | none stated |
| Camburn 1995 | X3 types Pre-contact or control | final follow-up | Randomised control trail. | Random digit dialling (USA). Non house hold and non-working numbers excluded | child immunisation |  | mailed | phone | none stated |
| Dillman 1976 | Pre-contact or control | only one mailing | Randomised control trail. | Random digit dialling (USA) |  |  | mailed | phone | none stated |
| Eyerman 2003 | Pre-contact or control | only one mailing | Randomised control trail. | Random digit dialling (USA) | Health risk factors | under a week | mailed | phone | none stated |
| Goldstein 2002 | Pre-contact or control | final follow-up | Randomised control trail. | Random digit dialling (USA) | Political polling | under a week | mailed | phone | none stated |
| Hembroff 2005 | X2 types Pre-contact or control | final follow-up | Randomised control trail. | Random digit dialling (USA) |  |  | Mailed | phone | none stated |
| Iredell 2005 | Pre-contact or control | final follow-up | Randomised control trail. | electoral roll. Have to be over 60 (Australia) | road crossing behaviour | 2 weeks | mailed | phone | none stated |
| Micky 1999 | (Pre-contact or control) x (survey administration or control) | final follow-up | factorial experiment | over 40 (USA) |  | one week | mailed | in person or phone | none stated |
| Smith 1995 | Pre-contact or control | only one mailing | Randomised control trail. | Random digit dialling (Australia). Exclude if non residential household, or non English speaking | health questions |  | mailed | phone | none stated |
| Singer 2000 | Pre-contact or control | only one mailing | Randomised control trail. | Random digit dialling (USA) | consumer attitudes |  | mailed | phone | none stated |
| Grritsenen 2002 A | Pre-contact or control | final follow-up | Randomised control trail. | Random digit dialling (Netherlands) | meat consumption, non-commercial | one week | mailed | phone | none stated |
| Grritsenen 2002 B | Pre-contact or control | final follow-up | Randomised control trail. | people who had not answered their phone | meat consumption, non-commercial | one week | answer phone message | phone | none stated |
| Brick 1997 | Pre-contact or control | final follow-up | Randomised control trail. | Random digit dialling (USA) |  |  | mailed | phone | Asked questions |
