## Supplemental Table 6 for "Does advance contact with research participants increase response to questionnaires: A Systematic Review and meta-Analysis"

Supplementary Table 3: Judgment and supporting evidence of study risk of bias.

| **Study** | **Sequence Generation: (Judgement) support** | **Allocation Concealment: (Judgement) support** | **Patients and Personal Blinding: (Judgement) support** | **Blinding of Outcome Assessment: (Judgement) support** | **Incomplete Outcome Data: (Judgement) support** | **Selective Reporting: (Judgement) support** | **Other Biases: (Judgement) support)** | **Overall Risk: (Judgement) support** |
| --- | --- | --- | --- | --- | --- | --- | --- | --- |
| Bergen 1957 | (unclear)  not specified | (unclear)  not mentioned | (low)  communication with study team pro forma, patients blind to randomisation | (low)  not mentioned; outcome objective | (low)  outcome data on all patients | (low)  no evidence of | (low)  no other bias apparent | (unclear)  unclear in some domains |
| Albaum 1989 | (unclear)  “… assigned randomly …” | (unclear)  not mentioned | (low)  communication with study team pro forma, patients blind to randomisation | (low)  not mentioned; outcome objective | (low)  outcome data on all patients | (low)  no evidence of | (low)  no other bias apparent | (unclear)  unclear in some domains |
| Drummond 2008 | (low)  “… random allocation using random number generator …” | (low)  confirmed in correspondence | (low)  communication with study team pro forma, patients blind to randomisation | (low)  not mentioned; outcome objective | (low)  outcome data on all patients | (low)  no evidence of | (low)  no other bias apparent | (low)  low in all domains |
| Napoles-Springer 2004 | (low)  “… randomly assigned using a random number table …” | (unclear)  not mentioned | (low)  communication with study team pro forma, patients blind to randomisation | (low)  not mentioned; outcome objective | (low)  outcome data on all patients | (low)  no evidence of | (low)  no other bias apparent | (unclear)  unclear in some domains |
| Newby 2003 | (low)  Confirmed in correspondence | (low)  Confirmed in correspondence | (low)  Confirmed in correspondence | (low)  not mentioned; outcome objective | (low)  outcome data on all patients | (low)  no evidence of | (low)  no other bias apparent | (low)  low in all domains |
| Ogborne 1986 | (unclear)  “… randomly assigned …” | (unclear)  not mentioned | (unclear)  study team blinded not stated, patients unaware of randomisation | (low)  not mentioned; outcome objective | (low)  outcome data on all patients | (low)  no evidence of | (low)  no other bias apparent | (unclear)  unclear in some domains |
| Whiteman 2003 | (low)  Confirmed in correspondence | (low)  confirmed in correspondence | (low)  communication with study team pro forma, patients blind to randomisation | (low)  not mentioned; outcome objective | (low)  outcome data on all patients | (low)  no evidence of | (low)  no interaction or other bias | (low)  low in all domains |
| Cycyota 2002 | (unclear)  “… a random half …” | (unclear)  not mentioned | (low)  communication with study team pro forma, patients blind to randomisation | (low)  not mentioned; outcome objective | (low)  outcome data on all patients | (low)  no evidence of | (high)  interaction with follow up | (high)  risk of bias from interaction |
| Childers 1979 | (unclear)  “… partitioned randomly …” | (unclear)  not mentioned | (low)  communication with study team pro forma, patients blind to randomisation | (low)  not mentioned; outcome objective | (low)  outcome data on all patients | (low)  no evidence of | (low)  no other bias apparent | (unclear)  unclear in some domains |
| Eaker 1998 | (unclear)  “factors were varied according to a randomised (unclear) (high) factorial design” | (unclear)  not mentioned | (low)  communication with study team pro forma, patients blind to randomisation | (low)  not mentioned; outcome objective | (low)  outcome data on all patients | (low)  no evidence of | (low)  no other bias apparent | (unclear)  unclear in some domains |
| Etter 1998 | (low)  “… list of random numbers sequenced by a computer (epi-info)” | (low)  confirmed in correspondence | (low)  communication with study team pro forma, patients blind to randomisation | (low)  not mentioned; outcome objective | (low)  outcome data on all patients | (low)  no evidence of | (low)  no other bias apparent | (low)  low in all domains |
| Ford 1967 A | (unclear)  “the no advance letter/advance letter process was random” (email correspondence) | (unclear)  not mentioned | (low)  communication with study team pro forma, patients blind to randomisation | (low)  not mentioned; outcome objective | (low)  outcome data on all patients | (low)  no evidence of | (low)  no other bias apparent | (unclear)  unclear in some domains |
| Ford 1967 B | (unclear)  “the no advance letter/advance letter process was random” (email correspondence) | (unclear)  not mentioned | (low)  communication with study team pro forma, patients blind to randomisation | (low)  not mentioned; outcome objective | (low)  outcome data on all patients | (low)  no evidence of | (low)  no other bias apparent | (unclear)  unclear in some domains |
| Hansen 1980 | (unclear)  Sent to a “… random group …” | (unclear)  not mentioned | (unclear)  study team blinded not stated, patients unaware of randomisation | (low)  not mentioned; outcome objective | (low)  outcome data on all patients | (low)  no evidence of | (low)  no interaction or other bias | (unclear)  unclear in some domains |
| Harrison 2004 | (low)  “… randomisation was done using computer generated random numbers ...” | (low)  Confirmed in correspondence | (low)  communication with study team pro forma, patients blind to randomisation | (low)  not mentioned; outcome objective | (low)  outcome data on all patients | (low)  no evidence of | (low)  no other bias apparent | (low)  low in all domains |
| Hornik 1982 | (unclear)  “… assigned randomly …” | (unclear)  not mentioned | (unclear)  study team blinded not stated, patients unaware of randomisation | (low)  not mentioned; outcome objective | (low)  outcome data on all patients | (low)  no evidence of | (low)  no other bias apparent | (unclear)  unclear in some domains |
| Kephart 1958 | (unclear)  “… randomly divided …” | (unclear)  not mentioned | (low)  communication with study team pro forma, patients blind to randomisation | (low)  not mentioned; outcome objective | (low)  outcome data on all patients | (low)  no evidence of | (low)  no interaction or other bias | (unclear)  unclear in some domains |
| Mann 2005 | (low)  communication: SPSS | (low)  communication: allocation automated | (low)  communication with study team pro forma, patients blind to randomisation | (low)  not mentioned; outcome objective | (low)  outcome data on all patients | (low)  no evidence of | (low)  no other bias apparent | (low)  low in all domains |
| Parsons 1972 A | (unclear)  sampled randomly with probabilities proportional to size of population | (low)  confirmed in correspondence | (low)  communication with study team pro forma, patients blind to randomisation | (low)  not mentioned; outcome objective | (low)  outcome data on all patients | (low)  no evidence of | (low)  no other bias apparent | (unclear)  unclear in some domains |
| Parsons 1972 B | (unclear)  sampled randomly with probabilities proportional to size of population | (low)  confirmed in correspondence | (low)  communication with study team pro forma, patients blind to randomisation | (low)  not mentioned; outcome objective | (low)  outcome data on all patients | (low)  no evidence of | (low)  no other bias apparent | (unclear)  unclear in some domains |
| Pirotta 1999 | (low)  “… sent randomly …” | (high)  communication: "probably nil" | (low)  communication with study team pro forma, patients blind to randomisation | (low)  “One person (MP), who was blinded to the intervention status … completed data entry” | (low)  outcome data on all patients | (low)  no evidence of | (low)  no other bias apparent | (high)  risk of bias from no allocation concealment |
| Shiono 1991 | (low)  “… randomised with equal probability, using a computer generated simple randomisation scheme …” | (unclear)  not mentioned | (low)  communication with study team pro forma, patients blind to randomisation | (low)  not mentioned; outcome objective | (low)  outcome data on all patients | (low)  no evidence of | (low)  no interaction or other bias | (unclear)  unclear in some domains |
| Spry 1989 | (low)  “… randomly assigned …” | (high)  confirmed in correspondence | (unclear)  communication with study team pro forma for letter, no blinding for call; patients blind to randomisation | (low)  not mentioned; outcome objective | (low)  outcome data on all patients | (low)  no evidence of | (low)  no other bias apparent | (high)  no allocation concealment |
| Wiseman 1972 | (unclear)  Split “on an experimental basis …” | (unclear)  not mentioned | (unclear)  study team blinded not stated, patients unaware of randomisation | (low)  not mentioned; outcome objective | (low)  outcome data on all patients | (low)  no evidence of | (low)  no other bias apparent | (unclear)  unclear in some domains |
| Wright 1995 | (high)  The list was ordered alphabetically and systematically sampled (three names at a time) | (low)  correspondence: Respondents in the control group were unaware of the presence of a pre-notification letter for the experimental group, and vice versa. | (low)  communication with study team pro forma, patients blind to randomisation | (low)  not mentioned; outcome objective | (low)  outcome data on all patients | (low)  no evidence of | (low)  no other bias apparent | (high)  High in one domain. |
| Dillman 1974 | (unclear)  Confirmed in correspondence | (unclear)  not mentioned | (unclear)  study team blinded not stated, patients unaware of randomisation | (low)  not mentioned; outcome objective | (low)  outcome data on all patients | (low)  no evidence of | (low)  no other bias apparent | (unclear)  unclear in some domains |
| Furst 1979 | (low)  “computer generated random number was used to allocate to group (email)” | (high)  no concealment – confirmed in correspondence | (low)  communication with study team pro forma, patients blind to randomisation | (low)  not mentioned; outcome objective | (low)  outcome data on all patients | (low)  no evidence of | (low)  no other bias apparent | (high)  risk of bias from allocation concealment |
| Gillpatick 1994 | (unclear)  systematic random sampling | (high)  no concealment – confirmed in correspondence | (low)  communication with study team pro forma, patients blind to randomisation | (low)  not mentioned; outcome objective | (low)  outcome data on all patients | (low)  no evidence of | (low)  no other bias apparent | (high)  risk of bias from allocation concealment |
| Heaton 1965 | (unclear)  “… randomly divided in half” | (unclear)  not mentioned | (low)  communication with study team pro forma, patients blind to randomisation | (low)  not mentioned; outcome objective | (low)  outcome data on all patients | (low)  no evidence of | (low)  no other bias apparent | (unclear)  unclear in some domains |
| Jobber 1985 | (unclear)  systematic random sampling (email correspondence) | (unclear)  not mentioned | (unclear)  study team blinded not stated, patients unaware of randomisation | (low)  not mentioned; outcome objective | (low)  outcome data on all patients | (low)  no evidence of | (low)  no other bias apparent | (unclear)  unclear in some domains |
| Jobber 1983 | (unclear)  “… randomly allocated…” | (unclear)  not mentioned | (low)  communication with study team pro forma, patients blind to randomisation | (low)  not mentioned; outcome objective | (low)  outcome data on all patients | (low)  no evidence of | (low)  no interaction or other bias | (unclear)  unclear in some domains |
| Kindra 1985 | (unclear)  not stated | (high)  inadequate in communication | (unclear)  study team blinded not stated, patients unaware of randomisation | (low)  not mentioned; outcome objective | (low)  outcome data on all patients | (low)  no evidence of | (low)  no interaction or other bias | (high)  risk of bias from allocation concealment |
| Myers 1969 | (unclear)  not stated | (unclear)  not mentioned | (low)  communication with study team pro forma, patients blind to randomisation | (low)  not mentioned; outcome objective | (low)  outcome data on all patients | (low)  no evidence of | (low)  no other bias apparent | (unclear)  unclear in some domains |
| Nichols 1988 | (unclear)  alternative member from systematic sample | (unclear)  not mentioned | (low)  communication with study team pro forma, patients blind to randomisation | (low)  not mentioned; outcome objective | (low)  outcome data on all patients | (low)  no evidence of | (low)  no other bias apparent | (unclear)  unclear in some domains |
| Osborn 1996 | (low)  Using “SAS statistical software” | (unclear)  not mentioned | (unclear)  study team blinded not stated, patients unaware of randomisation | (low)  not mentioned; outcome objective | (low)  outcome data on all patients | (low)  no evidence of | (low)  no other bias apparent | (unclear)  unclear in some domains |
| Pucel 1971 | (unclear)  “… randomly assigned to …” | (unclear)  not mentioned | (low)  communication with study team pro forma, patients blind to randomisation | (low)  not mentioned; outcome objective | (low)  outcome data on all patients | (low)  no evidence of | (low)  no interaction or other bias | (unclear)  unclear in some domains |
| Duhan 1990 | (unclear)  “… randomly assigned …” | (low)  communication None of the participants knew of their trail arm or of other trail arms. | (low)  communication with study team pro forma, patients blind to randomisation | (low)  not mentioned; outcome objective | (low)  outcome data on all patients | (low)  no evidence of | (low)  no other bias apparent | (unclear)  unclear in some domains |
| Faria 1990 | (low)  Each group sampled systematically from list. | (unclear)  not mentioned | (unclear)  communication with study team pro forma for letter, no blinding for call; patients blind to randomisation | (low)  not mentioned; outcome objective | (low)  outcome data on all patients | (low)  no evidence of | (low)  no other bias apparent | (unclear)  unclear in some domains |
| Stafford 1966 | (unclear)  “the total sample randomly divided into three segments” | (unclear)  not mentioned | (unclear)  communication with study team pro forma for letter, no blinding for call; patients blind to randomisation | (low)  not mentioned; outcome objective | (low)  outcome data on all patients | (low)  no evidence of | (low)  no other bias apparent | (unclear)  unclear in some domains |
| Sutton 1992 | (unclear)  “… randomly assigned …” | (low)  confirmed in correspondence | (low)  communication with study team pro forma, patients blind to randomisation | (low)  not mentioned; outcome objective | (low)  outcome data on all patients | (low)  no evidence of | (low)  no other bias apparent | (unclear)  unclear in some domains |
| Taylor 1998 | (unclear)  stratified random method | (unclear)  not mentioned | (low)  communication with study team pro forma, patients blind to randomisation | (low)  not mentioned; outcome objective | (low)  outcome data on all patients | (low)  no evidence of | (low)  no other bias apparent | (unclear)  unclear in some domains |
| Martin 1989 | (unclear)  “randomly assigned” | (unclear)  not mentioned | (low)  communication with study team pro forma, patients blind to randomisation | (low)  not mentioned; outcome objective | (low)  outcome data on all patients | (low)  no evidence of | (high)  two way interaction of per-contact and personalisation. | (high)  risk of bias from interaction |
| Chebat 1991 | (unclear)  not specified | (low)  confirmed in correspondence | (low)  communication with study team pro forma, patients blind to randomisation | (low)  not mentioned; outcome objective | (low)  outcome data on all patients | (low)  no evidence of | (low)  no other bias apparent | (unclear)  unclear in some domains |
| Xie 2013 | (low)  communication: "Used Excel to generate the random number" | (low)  random allocation was done by an independent research assistant | (low)  the investigators and participants remained unaware about the group allocation. | (low)  not mentioned; outcome objective | (low)  outcome data on all patients | (low)  no evidence of | (low)  no other bias apparent | (low)  low in all domains |
| Mitchell 2012 | (low)  “… a computer program randomly divided …” | (low)  “The allocation was independent and concealed” | (low)  communication with study team pro forma, patients blind to randomisation | (low)  not mentioned; outcome objective | (low)  outcome data on all patients | (low)  no evidence of | (low)  no other bias apparent | (low)  low in all domains |
| Maclennan 2014 | (low)  “Eligible participants were … randomised …” | (low)  confirmed in correspondence | (unclear)  study team blinded not stated, patients unaware of randomisation | (low)  not mentioned; outcome objective | (low)  outcome data on all patients | (low)  no evidence of | (low)  no other bias apparent | (unclear)  unclear in some domains |
| Keding 2016 | (low)  communication: used a computer programme using simple randomisation (data manager did the allocation). | (low)  randomisation by an independent data manager | (low)  “Participants did not know that they were part of an RCT … Text messaging was automated | (low)  not mentioned; outcome objective | (low)  outcome data on all patients | (low)  no evidence of | (low)  no other bias apparent | (low)  low in all domains |
| Hammink 2010 | (low)  communication: Computer-generated random numbers | (low)  random allocation done by independent statistician, who was not involved in the project | (low)  communication with study team pro forma, patients blind to randomisation | (low)  not mentioned; outcome objective | (low)  outcome data on all patients | (low)  no evidence of | (low)  no other bias apparent | (low)  low in all domains |
| Felix 2011 | (low)  “One author (L.F.) conducted the randomization using the RANDBETWEEN() command in Microsoft Excel.” | (low)  “At the time of randomization, none of the authors were aware of which intervention each group of participants would be allocated.” | (low)  communication with study team pro forma, patients blind to randomisation | (low)  The research team remained blind until data collection was complete | (low)  outcome data on all patients | (low)  no evidence of | (low)  *No evidence for interaction.* | (low)  low in all domains |
| Bauman 2016 | (low)  “Using the SAS statistical program, three thousand participants were randomly sampled … and allocated to three groups …“ | (low)  “statistician blinded to participant identity performed the randomisation sequence and was” otherwise independent | (low)  “logistics agency was contracted to administer the mail-put of questionnaire packages and staff were not informed of the allocation.” | (low)  not mentioned; outcome objective | (low)  outcome data on all patients | (low)  no evidence of | (low)  no other bias apparent | (low)  low in all domains |
| Barra 2016 | (low)  used 'random allocation'. correspondence: conducted by external statistician | (low)  Done by external independent statistician | (low)  communication with study team pro forma, patients blind to randomisation | (low)  not mentioned; outcome objective | (low)  outcome data on all patients | (low)  no evidence of | (low)  Pre-registered trail. Use ITT | (low)  low in all domains |
| Bosnjak 2008 | (low)  communication: used MS Excel | (low)  communication: the experimental variation was not communicated to study subjects. | (low)  communication with study team pro forma, patients blind to randomisation | (low)  not mentioned; outcome objective | (unclear)  Participants potentially excluded | (low)  no evidence of | (low)  no interaction or other bias | (unclear)  unclear in some domains |
| Boyd 2015 | (low)  Used “random number generator from STATA, ….” | (low)  “data was pseudonimized during allocation. State it is consolidate” | (low)  communication with study team pro forma, patients blind to randomisation | (low)  not mentioned; outcome objective | (low)  outcome data on all patients | (low)  no evidence of | (low)  no interaction or other bias | (low)  low in all domains |
| Dykema 2011 | (unclear)  “…randomly assigning …” | (low)  communication: Sample members were randomized into treatment groups and received personalized letters and emails that requested survey participation. Each mailing noted which incentive they were to receive, but made no mention of other treatment groups, or to the incentive experiment. No other concealment was deemed useful or necessary. | (low)  communication with study team pro forma, patients blind to randomisation | (low)  not mentioned; outcome objective | (low)  outcome data on all patients | (low)  no evidence of | (low)  no interaction or other bias | (unclear)  unclear in some domains |
| Grande 2016 | (unclear)  “… were randomly selected to be sent a SMS.” | (unclear)  not mentioned | (low)  communication with study team pro forma, patients blind to randomisation | (low)  not mentioned; outcome objective | (low)  outcome data on all patients | (low)  no evidence of | (low)  no other bias apparent | (unclear)  unclear in some domains |
| Keusch 2012 | (high)  communication: systematically based alphabetical order | (unclear)  not mentioned | (low)  communication with study team pro forma, patients blind to randomisation | (low)  not mentioned; outcome objective | (high)  363 exlcued because they could not be contacted | (low)  no evidence of | (low)  no interaction or other bias | (high)  High in at least one domain |
| Mclean 2014 | (low)  “.. randomly allocated … Using an Excel random number generator.” | (low)  communication: Random sequence generation was conducted independently by the first author; the sequence and allocation was then provided to the third author, who implemented the procedure for provision of the experimental conditions. | (low)  communication with study team pro forma, patients blind to randomisation | (low)  not mentioned; outcome objective | (low)  outcome data on all patients | (low)  no evidence of | (low)  no interaction or other bias | (low)  low in all domains |
| Rao 2010 | (unclear)  “… randomly divided …”. | (unclear)  not mentioned | (low)  communication with study team pro forma, patients blind to randomisation | (low)  not mentioned; outcome objective | (low)  outcome data on all patients | (low)  no evidence of | (low)  no interaction or other bias | (unclear)  unclear in some domains |
| Starr 2015 | (low)  “… randomly assigned …”. | (low)  was concealed and remote from the users | (low)  state none done; communication with study team pro forma, patients blind to randomisation | (low)  not mentioned; outcome objective | (low)  outcome data on all patients | (low)  no evidence of | (low)  no interaction | (low)  low in all domains |
| Veen 2015 | (unclear)  “… randomly assigned …” | (unclear)  not mentioned | (low)  communication with study team pro forma, patients blind to randomisation | (low)  not mentioned; outcome objective | (low)  outcome data on all patients | (low)  no evidence of | (low)  no other bias apparent | (unclear)  unclear in some domains |
| Ho-A-Yun 2007 | (low)  communication: Computer generated by an independent researcher | (unclear)  communication: The allocation was performed by Fay Crawford and the other investigators did not have access to the random allocation. I suspect the 2 groups were coded using 2 different colours of paper for the prompted versus not prompted group questionnaires but I can’t remember. | (low)  study team blinded (communication), pts unaware of randomisation | (low)  not mentioned; outcome objective | (low)  outcome data on all patients | (low)  no evidence of | (low)  no other bias apparent | (low)  low in all domains |
| Porter 2007 A | (low)  correspondence: " either a SAS sampling routine or the random number function in Excel" | (unclear)  not mentioned | (low)  communication with study team pro forma, patients blind to randomisation | (low)  not mentioned; outcome objective | (low)  outcome data on all patients | (low)  no evidence of | (low)  no interaction or other bias | (unclear)  unclear in some domains |
| Porter 2007 B | (low)  correspondence: " either a SAS sampling routine or the random number function in Excel" | (unclear)  not mentioned | (low)  communication with study team pro forma, patients blind to randomisation | (low)  not mentioned; outcome objective | (low)  outcome data on all patients | (low)  no evidence of | (low)  no interaction or other bias | (unclear)  unclear in some domains |
| Atinc 2012 | (low)  “randomisation … was handled by a computer system (Qualtrics)” | (low)  communication: The respondents were randomly assigned and they were not informed about their group | (low)  communication with study team pro forma, patients blind to randomisation | (low)  not mentioned; outcome objective | (low)  outcome data on all patients | (low)  no evidence of | (low)  no interaction or other bias | (low)  low in all domains |
| Walker 1977 | (unclear)  method of allocation not state | (unclear)  not mentioned | (low)  communication with study team pro forma, patients blind to randomisation | (low)  not mentioned; outcome objective | (low)  outcome data on all patients | (low)  no evidence of | (low)  no other bias apparent | (unclear)  unclear in some domains |
| Snow 1986 | (unclear)  “…randomly selected …” | (unclear)  not mentioned | (low)  communication with study team pro forma, patients blind to randomisation | (low)  interviewer blind allocation and outcome objective. | (low)  outcome data on all patients | (low)  no evidence of | (low)  no other bias apparent | (unclear)  unclear in some domains |
| Pitiyanuwat 1991 | (unclear)  not stated | (unclear)  not mentioned | (low)  communication with study team pro forma, patients blind to randomisation | (low)  not mentioned; outcome objective | (low)  outcome data on all patients | (low)  no evidence of | (low)  no interaction or other bias | (unclear)  unclear in some domains |
| Nicolaas 2015 | (unclear)  Used systematic random sample in each arm | (unclear)  not mentioned | (low)  communication with study team pro forma, patients blind to randomisation | (low)  not mentioned; outcome objective | (low)  outcome data on all patients | (low)  no evidence of | (low)  no interaction or other bias | (unclear)  unclear in some domains |
| Lynn 1998 | (high)  PPS with alternative allocation to each condition | (unclear)  not mentioned | (low)  communication with study team pro forma, patients blind to randomisation | (low)  not mentioned; outcome objective | (low)  outcome data on all patients | (low)  no evidence of | (low)  no other bias apparent | (high)  risk of bias from randomisation |
| Link 2005 | (unclear)  “… randomly assigned …” | (unclear)  not mentioned | (low)  communication with study team pro forma, patients blind to randomisation | (low)  “interviewers were blind to which cases … receive[d] advance letter[s]” | (low)  outcome data on all patients | (low)  no evidence of | (low)  no other bias apparent | (unclear)  unclear in some domains |
| Kulka 1981 | (unclear)  “… randomly pre-assigned …” | (unclear)  not mentioned | (low)  communication with study team pro forma, patients blind to randomisation | (low)  not mentioned; outcome objective | (low)  outcome data on all patients | (low)  no evidence of | (low)  no interaction or other bias | (unclear)  unclear in some domains |
| Kaplowitz 2004 | (unclear)  “… samples of students” for each condition | (Low)  Done by intendent/external office | (low)  communication with study team pro forma, patients blind to randomisation | (low)  not mentioned; outcome objective | (low)  outcome data on all patients | (low)  no evidence of | (high)  was interaction with pre-notification and follow-up | (high)  risk of bias from interaction |
| Groves 1987 | (unclear)  “… experimentally assigned …” | (unclear)  not mentioned | (low)  communication with study team pro forma, patients blind to randomisation | (low)  not mentioned; outcome objective | (low)  outcome data on all patients | (low)  no evidence of | (low)  no interaction or other bias | (unclear)  unclear in some domains |
| Furse 1981 | (unclear)  not stated | (unclear)  not mentioned | (unclear)  study team blinded not stated, patients unaware of randomisation | (low)  not mentioned; outcome objective | (low)  outcome data on all patients | (low)  no evidence of | (low)  no interaction or other bias | (unclear)  unclear in some domains |
| Chebatt 1993 | (unclear)  not stated | (unclear)  not mentioned | (low)  communication with study team pro forma, patients blind to randomisation | (low)  not mentioned; outcome objective | (unclear)  Participants potentially excluded | (low)  no evidence of | (low)  no other bias apparent | (unclear)  unclear in some domains |
| Boser 1990 | (unclear)  “… randomly assigned ...” | (unclear)  not mentioned | (low)  communication with study team pro forma, patients blind to randomisation | (low)  not mentioned; outcome objective | (low)  outcome data on all patients | (low)  no evidence of | (low)  no other bias apparent | (unclear)  unclear in some domains |
| Bergsten 1984 | (unclear)  “… a random half …” | (unclear)  not mentioned | (unclear)  study team blinded not stated, patients unaware of randomisation | (low)  not mentioned; outcome objective | (low)  outcome data on all patients | (low)  no evidence of | (low)  no other bias apparent | (unclear)  unclear in some domains |
| Baulne 2009 | (unclear)  “two groups were created at random” | (unclear)  not mentioned | (low)  communication with study team pro forma, patients blind to randomisation | (low)  not mentioned; outcome objective | (low)  outcome data on all patients | (low)  no evidence of | (low)  n.b. response rate is weighted to generalise to whole population; no other bias | (unclear)  unclear in some domains |
| Henri 2012 | (unclear)  “assigned the targeted firms randomly “ | (unclear)  not mentioned | (unclear)  communication with study team pro forma for letter, no blinding for call; patients blind to randomisation | (low)  not mentioned; outcome objective | (low)  outcome data on all patients | (low)  no evidence of | (low)  no interaction or other bias | (unclear)  unclear in some domains |
| Lalasz 2014 | (unclear)  “randomly-assigned“ | (unclear)  not mentioned | (low)  communication with study team pro forma, patients blind to randomisation | (low)  not mentioned; outcome objective | (low)  outcome data on all patients | (low)  no evidence of | (low)  no other bias apparent | (unclear)  unclear in some domains |
| Lippy 2011 | (unclear)  “ randomly split” | (unclear)  not mentioned | (low)  communication with study team pro forma, patients blind to randomisation | (low)  not mentioned; outcome objective | (low)  outcome data on all patients | (low)  no evidence of | (low)  no other bias apparent | (unclear)  unclear in some domains |
| Lusinchi 2007 | (low)  correspondence: used MS excel | (low)  confirmed in correspondence | (low)  communication with study team pro forma, patients blind to randomisation | (low)  not mentioned; outcome objective | (low)  outcome data on all patients | (low)  no evidence of | (low)  no other bias apparent | (low)  Low in all domains |
| Mcallster 2016 | (unclear)  “randomly assigned “ | (unclear)  not mentioned | (low)  communication with study team pro forma, patients blind to randomisation | (low)  not mentioned; outcome objective | (low)  outcome data on all patients | (low)  no evidence of | (low)  no other bias apparent | (unclear)  unclear in some domains |
| Miner 1986 | (unclear)  “randomly assigned “ | (unclear)  not mentioned | (unclear)  communication with study team pro forma for letter, no blinding for call; patients blind to randomisation | (low)  not mentioned; outcome objective | (low)  outcome data on all patients | (low)  no evidence of | (low)  no other bias apparent | (unclear)  unclear in some domains |
| Mitchell 2010 | (low)  communication: used a computer programme (data manager) with a single block that was equal to the size of the number of people being mailed out to. | (low)  communication: questionnaires were packed into brown paper envelopes with approximately halve having a newsletter. The two sets of envelopes would have been matched to a patients ID, so concealment was achieved because no one knew the characteristics of the patients being matched to the envelope. | (low)  communication with study team pro forma, patients blind to randomisation | (low)  not mentioned; outcome objective | (low)  outcome data on all patients | (low)  no evidence of | (low)  no interaction or other bias | (low)  Low in all domains |
| Steeh 2007 | (unclear)  “randomly assigned“ | (unclear)  not mentioned | (low)  communication with study team pro forma, patients blind to randomisation | (low)  not mentioned; outcome objective | (low)  outcome data on all patients | (low)  no evidence of | (low)  no other bias apparent | (unclear)  unclear in some domains |
| Vogl 2018 | (unclear)  “randomly assigned” | (unclear)  not mentioned | (low)  communication with study team pro forma, patients blind to randomisation | (low)  not mentioned; outcome objective | (low)  outcome data on all patients | (low)  no evidence of | (low)  no other bias apparent | (unclear)  unclear in some domains |
| Woodruf 2006 | (unclear)  “randomly assigned” | (unclear)  not mentioned | (low)  communication with study team pro forma, patients blind to randomisation | (low)  “interviewers were unaware of whether the respondent had received a letter…” | (low)  outcome data on all patients | (low)  no evidence of | (low)  no other bias apparent | (unclear)  unclear in some domains |
| Traugott 1993 | (unclear)  not stated | (unclear)  not mentioned | (low)  communication with study team pro forma, patients blind to randomisation | (low)  not mentioned; outcome objective | (low)  outcome data on all patients | (low)  no evidence of | (low)  no other bias apparent | (unclear)  unclear in some domains |
| Traugott 1987 | (unclear)  not stated | (unclear)  not mentioned | (low)  communication with study team pro forma, patients blind to randomisation | (low)  not mentioned; outcome objective | (low)  outcome data on all patients | (low)  no evidence of | (low)  no interaction or other bias | (unclear)  unclear in some domains |
| Brehm 1994 | (unclear)  “randomly distributed“ | (unclear)  not mentioned | (low)  communication with study team pro forma, patients blind to randomisation | (low)  not mentioned; outcome objective | (low)  outcome data on all patients | (low)  no evidence of | (low)  no other bias apparent | (unclear)  unclear in some domains |
| Camburn 1995 | (unclear)  “assigned randomly” | (unclear)  not mentioned | (low)  communication with study team pro forma, patients blind to randomisation | (low)  not mentioned; outcome objective | (unclear)  Participants potentially excluded | (low)  no evidence of | (low)  no other bias apparent | (unclear)  unclear in some domains |
| Dillman 1976 | (unclear)  “randomly assigned” | (unclear)  not mentioned | (low)  communication with study team pro forma, patients blind to randomisation | (low)  not mentioned; outcome objective | (unclear)  Participants potentially excluded | (low)  no evidence of | (low)  no other bias apparent | (unclear)  unclear in some domains |
| Eyerman 2003 | (unclear)  “randomly assigned” | (unclear)  not mentioned | (low)  communication with study team pro forma, patients blind to randomisation | (low)  not mentioned; outcome objective | (low)  outcome data on all patients | (low)  no evidence of | (low)  no other bias apparent | (unclear)  unclear in some domains |
| Goldstein 2002 | (unclear)  “a random half” | (unclear)  not mentioned | (low)  communication with study team pro forma, patients blind to randomisation | (low)  “Interviewers … Were not aware as to which potential respondents had received the pre-notification.” | (low)  use ITT | (low)  no evidence of | (low)  no other bias apparent | (unclear)  unclear in some domains |
| Hembroff 2005 | (unclear)  “…were then also divided randomly” | (unclear)  not mentioned | (low)  communication with study team pro forma, patients blind to randomisation | (low)  not mentioned; outcome objective | (low)  outcome data on all patients | (low)  no evidence of | (low)  no other bias apparent | (unclear)  unclear in some domains |
| Iredell 2005 | (unclear)  “a random selection of participants were sent …” | (unclear)  not mentioned | (low)  communication with study team pro forma, patients blind to randomisation | (low)  not mentioned; outcome objective | (low)  outcome data on all patients | (low)  no evidence of | (low)  no other bias apparent | (unclear)  unclear in some domains |
| Micky 1999 | (unclear)  “ randomly selected and assigned” | (unclear)  not mentioned | (low)  communication with study team pro forma, patients blind to randomisation | (low)  not mentioned; outcome objective | (low)  outcome data on all patients | (low)  no evidence of | (low)  no interaction or other bias | (unclear)  unclear in some domains |
| Smith 1995 | (unclear)  “half were randomly allocated” | (unclear)  not mentioned | (low)  communication with study team pro forma, patients blind to randomisation | (low)  interviewers were blinded to weather the household received a letter | (low)  outcome data on all patients | (low)  no evidence of | (low)  no other bias apparent | (unclear)  unclear in some domains |
| Singer 2000 | (unclear)  “a random half” | (unclear)  not mentioned | (low)  communication with study team pro forma, patients blind to randomisation | (low)  Interviewers were blind to whether or not a letter had been sent | (low)  outcome data on all patients | (low)  no evidence of | (low)  no other bias apparent | (unclear)  unclear in some domains |
| Grritsenen 2002 A | (unclear)  not stated but chosen randomly | (unclear)  not mentioned | (low)  communication with study team pro forma, patients blind to randomisation | (low)  not mentioned; outcome objective | (low)  outcome data on all patients | (low)  no evidence of | (low)  no other bias apparent | (unclear)  unclear in some domains |
| Grritsenen 2002 B | (unclear)  not stated but chosen randomly | (unclear)  not mentioned | (low)  communication with study team pro forma, patients blind to randomisation | (low)  not mentioned; outcome objective | (low)  outcome data on all patients | (low)  no evidence of | (low)  no other bias apparent | (unclear)  unclear in some domains |
| Brick 1997 | (low)  communication: Samples were assigned random numbers using SAS random number generator with a pre-specified initial start. | (low)  communication: The random numbers were assigned to all sample cases prior to any work so the staff had no recourse to the allocation. | (low)  communication with study team pro-forma, patients blind to randomisation | (low)  not mentioned; outcome objective | (low)  outcome data on all patients | (low)  no evidence of | (low)  no other bias apparent | (low)  Low in all domains |
