## Supplemental Table 7 for "Does advance contact with research participants increase response to questionnaires: A Systematic Review and meta-Analysis"

Supplementary Table 7: Search Terms

The following search terms were used to search in the databases stated above for potential studies, where ‘*’ represents truncation and ‘or’/’and’ represent the appropriate Boolean operators for the respective data base. No constraints were applied other than the stated time restrictions.

1. questionnaire
2. survey*
3. data collection
4. 1 or 2 or 3
5. respon*
6. return*
7. cooperat*
8. feedback
9. 5 or 6 or 7 or 8
10. control*
11. randomi*
12. blind*
13. mask*
14. trial*
15. compar*
16. experiment*
17. factorial
18. manipulation
19. 10 or 11 or 12 or 13 or 14 or 15 or 16 or 17 or 18
20. advance
21. advanced
22. earlier
23. prior
24. before*
25. already
26. afore
27. pre
28. previous
29. preceding
30. preliminary
31. introductory
32. primary
33. primer
34. foot-in-the-door
35. prompt*
36. prenotification
37. 20 or 21 or 22 or 23 or 24 or 25 or 26 or 27 or 28 or 29 or 30 or 31 or 32 or 33 or 34 or 35 or 36
38. letter
39. letters
40. postal
41. postcard
42. telephon*
43. telefon*
44. email*
45. mail*
46. leaflet*
47. phone
48. notice
49. remind*
50. communic*
51. contact
52. contacting
53. notification
54. foot-in-the-door
55. 38 or 39 or 40 or 41 or 42 or 43 or 44 or 45 or 46 or 47 or 48 or 49 or 50 or 51 or 52 or 53 or 54
56. 4 and 9 and 19 and 37 and 55
